# Machine learning approaches classify clinical malaria outcomes based on haematological parameters

**DOI:** 10.1101/2020.09.23.20200220

**Authors:** Collins M. Morang’a, Lucas Amenga–Etego, Saikou Y. Bah, Vincent Appiah, Dominic S. Amuzu, Nicholas Amoako, James Abugri, Abraham R. Oduro, Aubrey J. Cunnington, Gordon A. Awandare, Thomas D. Otto

**Affiliations:** West African Centre for Cell Biology of Infectious Pathogens (WACCBIP), University of Ghana, Accra, Ghana; Florey Institute, Molecular Biology and Biotechnology, University of Sheffield, Sheffield, UK; Department of Applied Chemistry and Biochemistry, C.K Tedam University of Technology and Applied Sciences, Navrongo, Ghana; Navrongo Health Research Centre (NHRC), Navrongo, Ghana; Section of Pediatric Infectious Disease, Department of Infectious Disease, Imperial College London, UK; Institute of Infection, Immunity & Inflammation, MVLS, University of Glasgow, Glasgow, UK

**Keywords:** Machine Learning, Uncomplicated & Severe Malaria, Classification

## Abstract

**Background:** Malaria is still a major global health burden, with more than 3.2 billion people in 91 countries remaining at risk of the disease. Accurately distinguishing malaria from other diseases, especially uncomplicated malaria (UM) from non-malarial infections (nMI) remains a challenge. Furthermore, the success of rapid diagnostic tests (RDT) is threatened by *Pfhrp2/3* deletions and decreased sensitivity at low parasitemia. Analysis of haematological indices can be used to support identification of possible malaria cases for further diagnosis, especially in travelers returning from endemic areas. As a new application for precision medicine, we aimed to evaluate machine learning (ML) approaches that can accurately classify nMI, UM and severe malaria (SM) using haematological parameters.

**Methods:** We obtained haematological data from 2,207 participants collected in Ghana; nMI (n=978), UM (n=526), and SM (n=703). Six different machine learning approaches were tested, to select the best approach. An artificial neural network (ANN) with three hidden layers was used for multi-classification of UM, SM, and uMI. Binary classifiers were developed to further identify the parameters that can distinguish UM or SM from nMI. Local interpretable model-agonistic explanations (LIME) were used to explain the binary classifiers.

**Results:** The multi-classification model had greater than 85 % training and testing accuracy to distinguish clinical malaria from nMI. To distinguish UM from nMI, our approach identified platelet counts, red blood cell (RBC) counts, lymphocyte counts and percentages as the top classifiers of UM with 0.801 test accuracy (AUC = 0.866 and F1-score = 0.747). To distinguish SM from nMI, the classifier had a test accuracy of 0.960 (AUC= 0.983, and F1-score = 0.944) with mean platelet volume and mean cell volume being the unique classifiers of SM. Random forest was used to confirm the classifications and it showed that platelet and RBC counts were the major classifiers of UM, regardless of possible confounders such as patient age and sampling location.

**Conclusions:** The study provides proof of concept methods that classify UM and SM from nMI, showing that ML approach is a feasible tool for clinical decision support. In the future, ML approaches could be incorporated into clinical decision-support algorithms for the diagnosis of acute febrile illness, and monitoring response to acute SM treatment particularly in endemic settings.

## Background

In 2018, there were 228 million cases of malaria worldwide, 93% of which occurred in the African region [1]. Furthermore, approximately 450,000 deaths were reported, of which 61 % were children under 5 years old [1]. According to WHO 2018 report, over 2.7 billion US dollars were spent towards various control and elimination efforts to address the global burden of malaria [1]. This includes over 2.74 billion doses of artemisinin based combination therapies, procured in 2017 [1]. Unfortunately, incorrect diagnosis leads to incorrect treatment. It can increase the chances of antimalarial drug resistance, or for false negative diagnosis, it may result in misdiagnosis of malaria, appropriate treatment and progress to severe disease or death [2–4]. The gold standard for malaria diagnosis is microscopy, which requires extensive training, but rapid diagnostic tests (RDTs) have become the frontline diagnostic tools for malaria because of their ease of use at point-of-care [5].

One drawback of RDTs is the emergence of gene deletions of the target antigen, histidine rich protein (*Pfhrp2/3*), in the parasite genome, which render parasites undetectable by the most common RDTs [6]. Other challenges include insufficient sensitivity to detect low-level parasitemia, and the number of tests which need to be performed per positive result in settings with declining or low transmission [7–9]. Different problems are faced in non-endemic countries, where imported malaria must be suspected as a possible cause of fever before an RDT or microscopy would be performed in the first place, and failure to identify cases at first contact with health services often results in worse clinical outcomes [10, 11]. Therefore, improved and complementary malaria diagnostics techniques are required, which can overcome some or all of these limitations.

Complete blood counts (CBCs) is the most commonly performed laboratory test in most hospitals in both developing and developed countries. The CBC is usually relied upon to provide clues for diagnosis of patients where advanced methods for detection of specific diseases are lacking, with a parameter such as decreased platelet counts often associated with severe malaria (SM) [12, 13]. In addition, hemoglobin levels (Hb) are very important for the classification of SM cases [14]. Indeed, the changes in haematological parameters during clinical malaria have been studied extensively, to aid in the understanding of disease pathogenesis [15–21]. However, the diagnostic value of haematological parameters measured by commonly available automated haematology analyzers has not been fully studied using unbiased approaches such as ML techniques. These haematological parameters have the potential to be used in differentiating clinical malaria from other febrile illnesses, especially in areas where the reliability of RDTs is challenged by high prevalence of *Pfhrp2/3* deletion mutant parasites.

The potential and diagnostic value of all the CBC parameters for diagnosis of malaria can be realized by using machine learning (ML). ML approaches use algorithms based on statistical assumptions and mathematical rules to learn patterns and produce meaningful classifications based on the association of each variable with the disease outcome [22–26]. These classifications can then be applied to new disease cases to make classifications on the most probable cause. This classification capability of ML has not been extensively implemented in the diagnosis of clinical malaria.

To date, only a single study has reported the use of ML to diagnose malaria using clinical history and symptoms captured verbally and visually [27]. The sample size (n=376) was very small to deduce meaningful classifications and the author concluded that more work would be needed [27]. Despite this, there have been far reaching studies on application of ML in other areas of malaria research [28–32]. The diagnosis of malaria using ML on clinical datasets has been impaired by lack of large data, as well as difficulty in data curation. Moreover, classical modelling is prone to over-fitting or under-fitting of data [33], but recent approaches such as imputation, encoding, centering and scaling of variables, and model optimization [26] enable augmented use of ML in malaria classification.

We hypothesized that we can classify clinical malaria and non-malarial infections (nMI) with an ML approach. We first collected and curated data from 2,207 patients including (nMI) (n=978), uncomplicated malaria (UM) (n=703), and severe malaria (SM) (n=526). We generated ML models to classify clinical malaria (UM and SM) from nMI using haematological parameters.

## Methods

### Study population and sample collection

Standards for Reporting Diagnostic Accuracy Studies (STARD) guidelines [34, 35] were followed in this study. The current study utilizes unpublished data of 868 patients from a previous case-control study of SM conducted by the Navrongo Health Research Centre (NHRC) located in the Kassena-Nankana Districts (KNDs) in the Upper East Region of Northern Ghana. In the original study, children with acute febrile symptoms admitted to the Navrongo War Memorial Hospital (NWMH), the only referral facility in the KNDs were evaluated for inclusion into the study from August to December 2002 and May 2003 to April 2004. Full details of the study procedure, inclusion criteria, demographic and clinical characteristics of SM cases may be reviewed in Oduro et al [36].

In brief, inclusion criteria for SM cases was: (1); all children between 6-59 months who had fever (or history of fever in past 24 hours) and were admitted to the (NWMH), (2) residence in the Navrongo Health and Demographic Surveillance System area [36], and (3) willingness of parents/caregivers to offer informed consent. Criteria for SM diagnosis and enrollment into the original study were classified as having SM by the WHO standard guidelines, that include hemoglobin < 5g/dl or hematocrit < 15% [36, 37]. Ethical approval for the SM study was obtained from Navrongo Health Research Center (NHRC) Institutional Review Board (IRB), Noguchi Memorial Institute of Medical Research (NMIMR) IRB, Naval Medical Research Center IRB, and the Ghana Health Service (GHS) Ethics Committee. Informed consent was obtained and documented, followed by administration of a questionnaire about the presenting symptoms and clinical examinations. Participants who did not consent, meet the study inclusion criteria and those who had reported taking antimalarial treatment in the past two weeks were excluded from the study, while those who turned out to be malaria negative by standard microscopy were withdrawn from the study. All study samples were taken prior to initiation of treatment except for samples taken for clinical monitoring during admission or for the follow-up after discharge from hospital.

The nMI and UM participants were recruited in a hospital-based cross-sectional study involving two hospitals; Kintampo North-Municipal Hospital, Kintampo and Ledzokuku Krowor Municipal Assembly Hospital (LEKMA), Teshie in Accra. The inclusion criteria were; (1) outpatient children 1-15 years old, (2) presenting with fever or history of fever in the past 24 hours or axillary temperature ≥ 38°C, (3) and (4) signed informed consent by self (adolescents) and parent/guardian. The exclusion criterion was participants with known chronic disease or history of antimalarial drug use in the past two weeks. Ethical approval was also obtained from NMIMR, GHS, and Kintampo Health Research Centre (KHRC). A case was defined as nMI if the individual presenting to the hospital was malaria negative by RDTs and microscopy. Clinical data such as age, sex, body temperature and symptoms such as fever were collected on recruitment.

### Sample Collection Procedure

Venous blood was collected in the ante-cubital fossa. Tourniquet was not applied beyond one minute during venesection to avoid haemo-concentration, which could give erroneous results for all parameters measured. Samples were taken mostly between 8 am and 12 pm to avoid variations due to individuals’ activity (such as rehydration and food intake). Samples (5 mL) were taken into K3 EDTA tubes (BD Vacutainer; Becton Dickinson, NJ, USA). Samples that could not get analyzed within two hours from the time of collection were stabilized at 2-8°C to avoid changes that could occur in some haematological parameters should the sample be left on the bench for more than three hours. Samples were analysed not later than 24 hours from the time of sample storage at 2-8°C. No capillary blood sample was taken during the study as it presents with subtle variations from venous blood parameters. CBC analysis was performed using the automated ABX Micros 60 haematology analyzer. Data were manually cross-referenced twice for accuracy to ensure consistency in sample collection procedures.

### Statistical classifier: median split

Kernel density estimation, which is a non-parametric technique, was used to estimate the probability density function of each haematological parameter and kernel distribution for each parameter between nMI, UM and SM and visualized using density plots in R (R-version 4.0.2). Median value within each diagnostic group (nMI, UM, and SM) was computed, and the mean of any two group medians was used for ‘median split’ to generate a dichotomous variable for each parameter (low and high levels representing below and above median respectively)[38]. Contingency tables were used to summarize the relationship between clinical diagnosis (nMI, UM, and SM), and each dichotomous parameter. The general linear models for predictive analysis were used to explain the relationship between the clinical diagnosis and the dichotomous parameter. Odds ratios were computed through the exponent of the regression coefficients (logits) to estimate the strength of the relationship. Any OR with 95% confidence interval (CI) that includes a null value (1.0) indicated that the parameter was not significantly associated with clinical diagnosis. ANOVA was used to compare the model with null model and Chi-square test used to compute the degree of significance. All the analysis was done in R (R-version 4.0.2)

### Data pre-processing and normalization

A multivariate imputation via chained equations (MICE) plot was used to visualize the missing observations in the data. It was difficult to determine whether the missing values were missing ‘completely at random’, or ‘missing at random’ or ‘not at random’ to enable selection of imputation method. Therefore, the demographic/clinical data and microscopy results were not imputed and were not used for modeling. The majority of the haematological parameters had less than 5 % missing data and the missing values were imputed using MICE package in R. Each variable in the training and test data were transformed using Yeo-Johnson function, centered to have a mean of zero, and scaled to have a standard deviation of one. The original dataset (before pre-processing and normalization) is available in Additional File 1.

### Machine learning

Six ML algorithms were evaluated to identify the best algorithm that can classify the binary data. These include partial least squares (PLS) logistic regression, multiple adaptive regression splines (MARS), random forest, decision trees, support vector machine and artificial neural networks. PLS logistic regression was implemented by reducing the dimension of haematological parameters so as to increase accuracy. We used 10-fold cross-validation while tuning through 16 principal components (PC), whereby the optimal model used 2 PC. The optimal hyperparameters for MARS (with cross validation) were determined in a grid search of 30 different combinations of 3^rd^ degree and sampling 1000 terms to retain the final model [40]. Decision tree were implanted with the *rpart* function, which performs auto tuning with an optimal subtree of 10 total tree splits. Random forest and support vector machines were implemented by first performing a grid search to identify the optimal hyperparameters followed by classification analysis. Three ANN were developed, one multi-classification ANN (nMI vs UM vs SM) and two binary classifications denoted as ANN (UM and nMI), and ANN (SM and nMI). For each ANN, the data were split into 80 % training and 20 % testing. The outcome was the clinical diagnosis of the participant (as concluded by the attending clinicians) having either UM or nMI or SM. Haemoglobin and hematocrit levels were not included in the modeling because they are used to support diagnosis of malaria [12, 21, 37, 39]

### Hyperparameter tuning for artificial neural networks

The ANN was composed of an input layer of 15 haematological parameters. The loss was computed using categorical cross-entropy for the multi-classifier and binary cross-entropy for binary classifiers, while accuracy was used as the main evaluation metric. During training, the 80 % training data was further split into 70 % training and 30 % validation with randomization (Figure 1). Tensor board visualizations were used to check the dynamic graphs of our training and test metrics (Supplementary figure 2). Hyperparameters were tuned to identify the optimal model parameters for each classification. A hyper-grid was developed that adjusts the model capacity, normalization term, kernel regularization, and learning rate. To maximize the validation error performance, we tuned 12, 32, 64, 128, 256, 512 rectified linear units (ReLU) in three hidden layers. We used batch normalization on each hidden layer for gradient propagation and performance improvement. We varied the dropout rate from 0.1, 0.2, 0.3, and 0.4 in all the three layers to identify the best dropout regularization that prevents the model from latching to happenstance patterns that are not significant. We used “Adam” as the optimizer, but we varied the learning rate (0.1, 0.05, 0.001, and 0.0001) to find a global minimum. The tfruns R package was used to implement the hyper-grid in R, using 500 epochs, batch size of 64 and validation split of 0.3. These Keras models composed were initialized for all the three models (supplementary figure 1) and the optimal model was selected.

**Figure 1.**
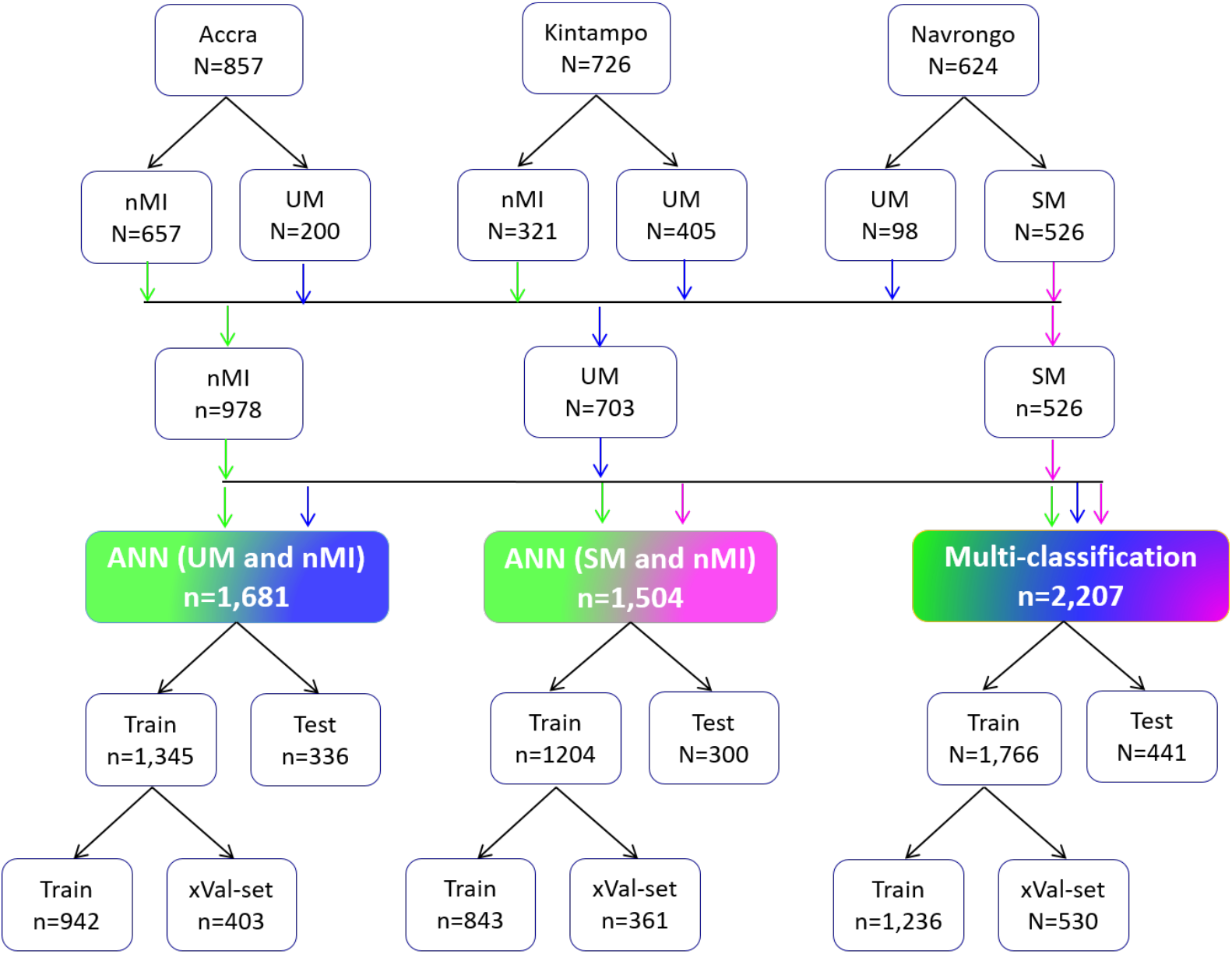
Study population and data splitting for building the ANN for clinical malaria. Samples were collected from two high transmission areas; Kintampo (n=726) and Navrongo (n=624), and one low transmission area (Accra, n = 857). The nMI (n = 978) were collected from Kintampo and Accra, UM (n = 703) were collected from all three areas, while the SM (n= 526) samples were collected from Navrongo. A multi-classification ANN model was developed for nMI, UM and SM, which was further evaluated by binary ANN models (1) ANN (UM vs nMI) and (2) ANN (SM vs nMI). For each model, data splitting was achieved by splitting data in 80:20% ratio into training (Train) and testing (Test). The 80% training data was further split into 70:30% ratio for training (Train) and cross-validation (xVal-set).

### ANN Model evaluations

Yardstick package was used to perform classifications on the test data as well as compute the performance of the model. The confusion matrix, accuracy, area under the Receiver Operating Characteristic Curve (AUC), precision and recall, and F1-Score were the metrics used to evaluate performance. The F1 score is a measure of test data accuracy, which is a weighted average between precision and recall. To explain the model, we used local interpretable model-agonistic explanations (LIME Package in R) [41]. The classification model was set up, and an “explainer” of the classifying model was developed using the training data and the model output classifications. The explainer was used to explain the results of the test dataset as classification explanations (feature weights). The feature weights were used to build a heatmap for each ANN indicating how each feature explains the model.

### Effect of patient age and sampling location on the model predictions

To test if patient age and sampling location significantly affected the models, we used three models (1) a model for all the UM and nMI cases (n=1,823), (2) a model for UM and nMI from Kintampo cases only (n=900), and (3) a model for only Kintampo cases and ages >4 years. We tested the possibility of using the ANN to evaluate the models but there was some level of over-fitting and under-fitting of the 2^nd^ and 3^rd^ models, due to sample size limitation. Therefore, random forest was subsequently used, because of (1) its robustness to smaller sample size with minimal over-fitting of the data (2) its ability to reduce the high variance from decision trees by combining several trees into one ensemble tree [42].

### Statistical analysis

The clinical categorical data was analyzed using Pearson’s Chi-square while the continuous data such as the haematological parameters were analyzed using Kruskal Wallis test with Dunn’s post hoc tests across the three groups (UM, SM and nMI). All tests were two sided and statistical significance was set at (*P<0*.*05*) for all analyses with adjustment for multiple testing. Data analyses was performed using R-development software (R version 4.0.2), R-studio (Version 1.1) and Python (Version 2.7). The R code with the methods, including the curated data files can be found on git-hub: https://github.com/misita-falcon.

## Results

### Characteristics of the study participants

Participants were recruited as follows; 38.8 % (857/2,207) from Accra, 32.9 % (726/2,207) from Kintampo, and 28.3 % (624/2,207) from Navrongo (Figure 1). These participants from all the three locations constitute 44.3 % (978/2,207) nMI, 31.8 % (703/2,207) for UM, and 23.8 % (526/2,207) for SM cases (Figure 1). The median age was 3 years (range: 2-6 years) for nMI, 4 years (range: 2-7 years) for UM, and 1 year (range: 1-2 years) for SM. The median ages were significantly different as determined by Kruskal Wallis test (*P*<.001) (Table 1). The sm cases had a significantly higher median body temperature (38.3; range =37.5-39.2), compared to the nMI (37.2; range = 36.5-38.4), UM (38.1; range = 37-39) and the SM cases (38.3; range =37.5-39.2) (*P*<.001). There was a significant difference in proportions of individuals (*P*<.001) among nMI, UM, and SM from different locations (Kintampo, Navrongo, and Accra) as determined by chi-square analysis (Table 1). There was no association between sex and clinical diagnosis, although the number of females was higher than males in all three groups (*P=*0.247); nMI was 51.2 % (501/978), UM was 54.9% (386/703) and SM was 55.1 % (290/526) (Table 1). Fever was more common in SM (99.2 %, 522/526) compared to UM (85.5 %, 601/703) and lowest in nMI (59.4%, 581/978), and chi-square analysis shows that there was an association between fever and clinical diagnosis (*P*<.001) (Table 1).

**Table 1.**
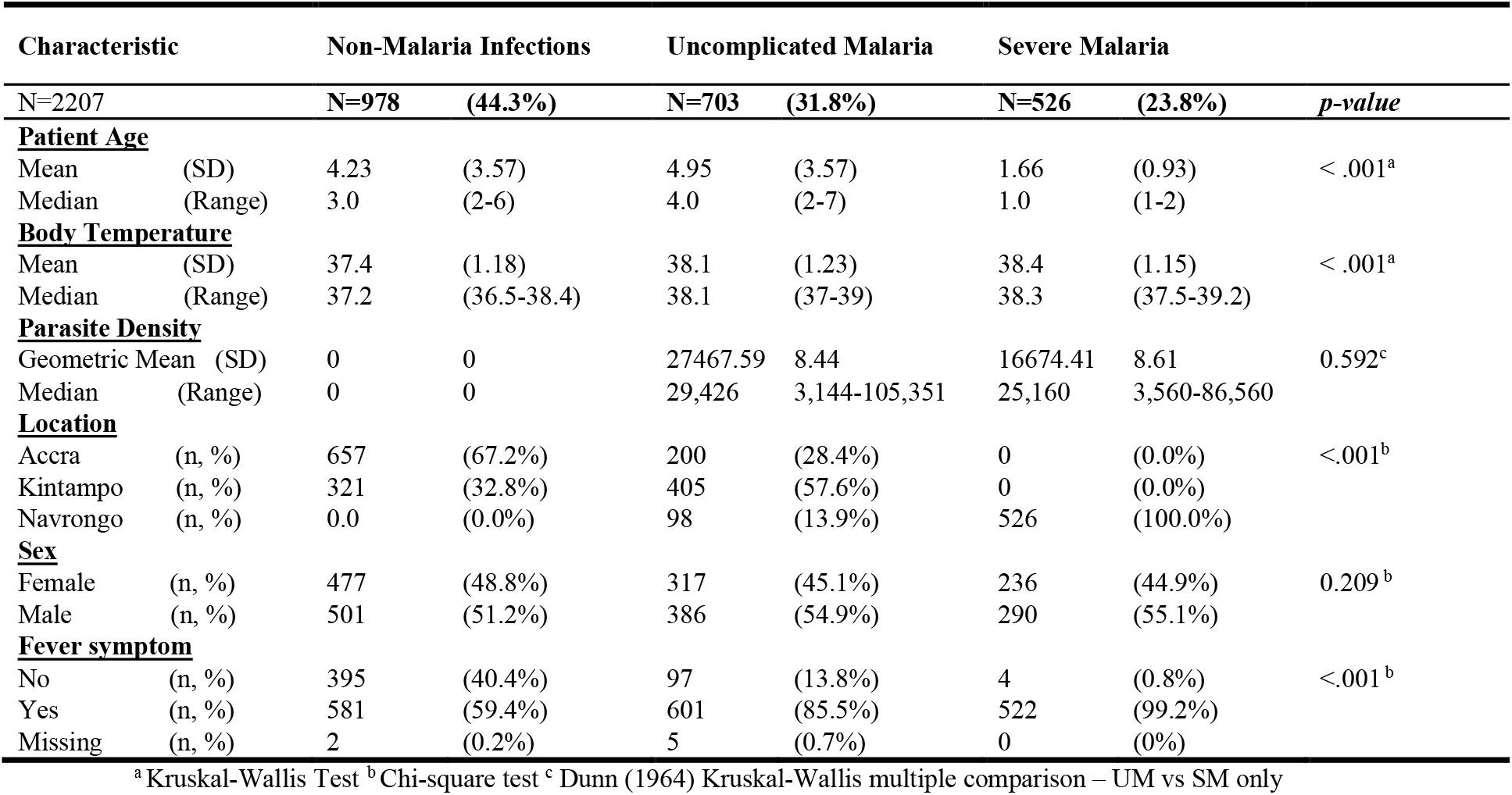
Characteristics of study participants for nMI, UM, an SM (n= 2,207). Average age and hemoglobin levels were analyzed using Kruskal Wallis test while recruitment location, sex and fever were analyzed using Chi-square test at 95% CI. All the participant characteristics were significantly different between the nMI, UM, and SM.

Participants with UM had a higher geometric mean density (27,467.59 Parasites/µL, SD=8.44) compared to SM individuals (16,674.41 Parasites/µL, SD=8.61). But, the median levels did not vary significantly between the two groups (*P*=0.592) (Table 1). Participants with nMI were negative by microscopy. There were 212 different suspected infections in the nMI group and the top 10 include; upper respiratory tract infections (17 %, 167/978), malaria (9.5 %, 93/978), gastroenteritis (7.6 %, 75/978), sepsis (6.1 %, 60/978), otitis media (5.9 %, 58/978), enteric fever (2.6%, 26/978), fever (2.1 %, 23/978), tonsillitis (2.3 %, 23/978), pneumonia (2.1%, 21/ 978) and anaemia (1.9 %, 19/978) (supplementary figure 1). Laboratory results indicated that majority of the samples were undetermined/not available/not known (96 %, 937/978), with only 4 % having accurate laboratory result (41/978). Some of the organisms that were laboratory confirmed include; *Streptococcus pneumonia, Staphylococcus aureus, Salmonelella thypi, Coxiella burnetii*, and Dengue virus (Figure 2). Only 2 UM participants had co-infections (laboratory confirmed) with *P. falciparum* and these individuals had *4* and *Group D streptococcus* (1). Since the sample size of laboratory confirmed nMI cases was low, all the samples were grouped as nMI, instead of individual diseases during ML classifications.

**Figure 2.**
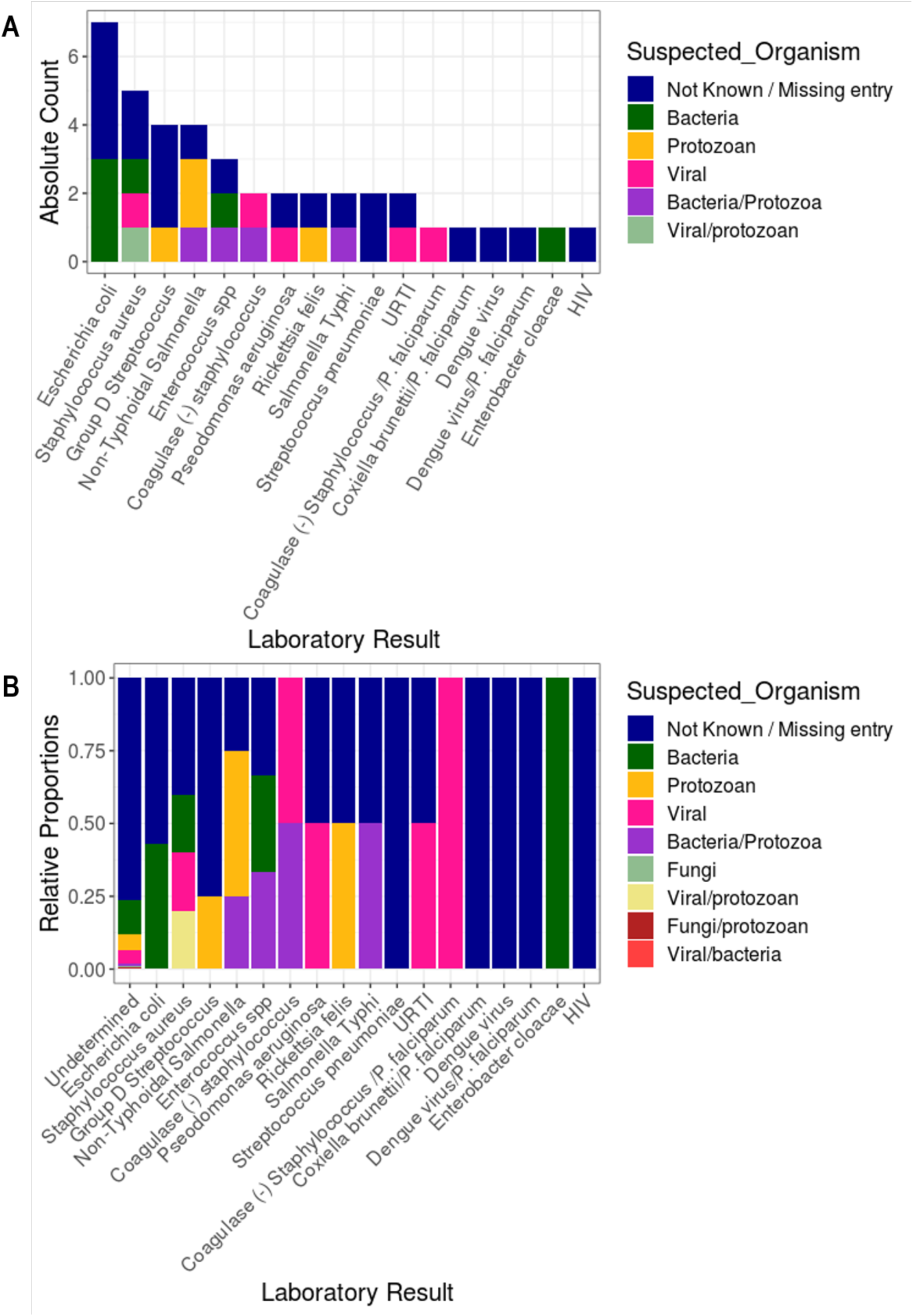
Clinical manifestations using laboratory diagnosis compared to various suspected infections by clinicians. Blood, urine and stool samples were collected from majority of the individuals who were categorized as nMI. Cultures of either blood, urine or stool were performed, depending on the clinicians/Doctor’s request and the suspected illness. The suspected organisms include bacteria, viruses and protozoan. Laboratory results confirmed only 4% of the cases with the majority being undetermined/not available/not known (96 %, 937/978). The major organisms determined to be present include: Dengue virus, *Staphylococcus aureus, Salmonella typhi, Streptococcus pneumonia*, and *Coxiella burnetii*.

### Haematological parameters vary between nMI, UM and SM

Median values for all the haematological parameters were significantly different among nMI, UM, and SM (*P*<.001) (Table 2), but most of the parameters do not show distinct distributions between the different clinical diagnosis groups (Figure 3). More so, Dunn’s post hoc tests indicated that platelet distribution width, percentage neutrophils and percentage lymphocytes were not significantly different between the nMI and SM (Table 2). Similarly, the pairwise comparisons showed that mean cell volume Neutrophils count and mean platelet volume were not significantly different between nMI and UM (Table 2). Despite the statistical test, we hypothesized that the median differences for each parameter cannot be used to confidently classify the disease outcomes.

**Table 2.** Comparison of median and interquartile ranges in haematology values measured in nMI, UM, and SM cases. *P-values* were analyzed using Kruskal Wallis test with post hoc tests (supplementary table 2). The parameters include WBC indices, RBC indices, and platelet indices. All the haematological parameters were significantly different between the nMI, UM, and SM (P<0.001) except the WBC counts, neutrophil counts, percent neutrophils and mean corpuscular hemoglobin.

|  | Non-Malaria Infections (a)<br>N=978 |  | Uncomplicated Malaria (b)<br>N=703 |  | *a vs b* | Severe Malaria (c)<br>N=526 |  | *b vs c* | *a vs c* | *a vs b vs c* |
| --- | --- | --- | --- | --- | --- | --- | --- | --- | --- | --- |
| Parameters | Media<br>n | IQR | Median | IQR | *P-value* | Median | IQR | *P-value* | *P-value* | *P-value* |
| **WBC Indices** |  |  |  |  |  |  |  |  |  |  |
| WBC Count ($10^3/\mu\text{L}$) | 9.3 | 7.0-12.8 | 8.3 | 6.3-10.8 | < .001 | 11.6 | 8.3-16.6 | < .001 | < .001 | <.001 |
| Lymphocytes Count ($10^3/\mu\text{L}$) | 3.0 | 2.0-4.5 | 1.9 | 1.3-3.0 | < .001 | 3.8 | 2.4-6.0 | < .001 | < .001 | <.001 |
| Mixed Cell Count ($10^3/\mu\text{L}$) | 0.8 | 0.5-1.1 | 0.5 | 0.3-0.8 | < .001 | 0.9 | 0.5-1.4 | < .001 | 0.004 | <.001 |
| Neutrophils Count ($10^3/\mu\text{L}$) | 4.8 | 3.3-7.6 | 5.4 | 3.7-7.6 | **0.115** | 6.5 | 4.4-9.4 | < .001 | < .001 | <.001 |
| Lymphocytes Percent (%) | 35.8 | 22.6-47.8 | 24.7 | 16.8-36.8 | < .001 | 33.9 | 26.5-44.4 | < .001 | **0.964** | <.001 |
| Mixed Cells Percent (%) | 8.6 | 6.7-11.0 | 6.9 | 5.0-9.2 | < .001 | 8.2 | 5.5-11.3 | < .001 | 0.012 | <.001 |
| Neutrophils Percent (%) | 54.4 | 41.7-69.0 | 67.8 | 53.7-77.1 | < .001 | 55.8 | 46.6-66.2 | < .001 | **0.568** | <.001 |
| **RBC Indices** |  |  |  |  |  |  |  |  |  |  |
| RBC Count ($10^6/\mu\text{L}$) | 4.5 | 4.2-5.0 | 4.1 | 3.6-4.5 | < .001 | 2.4 | 1.7-3.2 | < .001 | < .001 | <.001 |
| Hb Level (g/dL) | 11.0 | 10.1-11.8 | 10.1 | 8.8-11.2 | < .001 | 5.6 | 4.1-7.4 | < .001 | < .001 | <.001 |
| Hematocrit (%) | 34.5 | 32-37.1 | 31.1 | 27.2-34.8 | < .001 | 16.7 | 12.0-21.1 | < .001 | < .001 | <.001 |
| RBC Distribution Width (%) | 15.1 | 14.0-16.6 | 15.7 | 14.7-17.1 | < .001 | 18.1 | 16.2-20.1 | < .001 | < .001 | <.001 |
| Mean Cell Volume (fL) | 76.0 | 71.2-80.3 | 76.0 | 72.0-81.0 | **0.510** | 70.0 | 64.7-75.4 | < .001 | < .001 | <.001 |
| Mean Corpuscular Hb (pg) | 23.7 | 21.8-25.6 | 24.9 | 23.0-26.4 | < .001 | 23.8 | 21.5-26.7 | 0.001 | 0.006 | <.001 |
| Mean Cell Hb Concentration (g/dL) | 31.6 | 29.6-32.5 | 32.3 | 31.5-33.3 | < .001 | 35.1 | 31.2-37.4 | < .001 | < .001 | <.001 |
| **Platelet Indices** |  |  |  |  |  |  |  |  |  |  |
| Platelet Count ($10^3/\mu\text{L}$) | 292.0 | 226.0-360.0 | 140.0 | 92.0-216.0 | < .001 | 98.0 | 61.0-156.0 | < .001 | < .001 | <.001 |
| Mean Platelet Volume (fL) | 8.2 | 7.6-8.9 | 8.1 | 7.5-8.9 | **0.186** | 6.9 | 6.4-7.8 | < .001 | < .001 | <.001 |
| Platelet Distribution Width (fL) | 15.0 | 13.9-15.4 | 14.5 | 12.4-15.6 | 0.005 | 15 | 12.0-17.3 | < .001 | **0.121** | <.001 |

*P-value* – Kruskal Wallis test with Dunn’s post hoc tests

**Figure 3:**
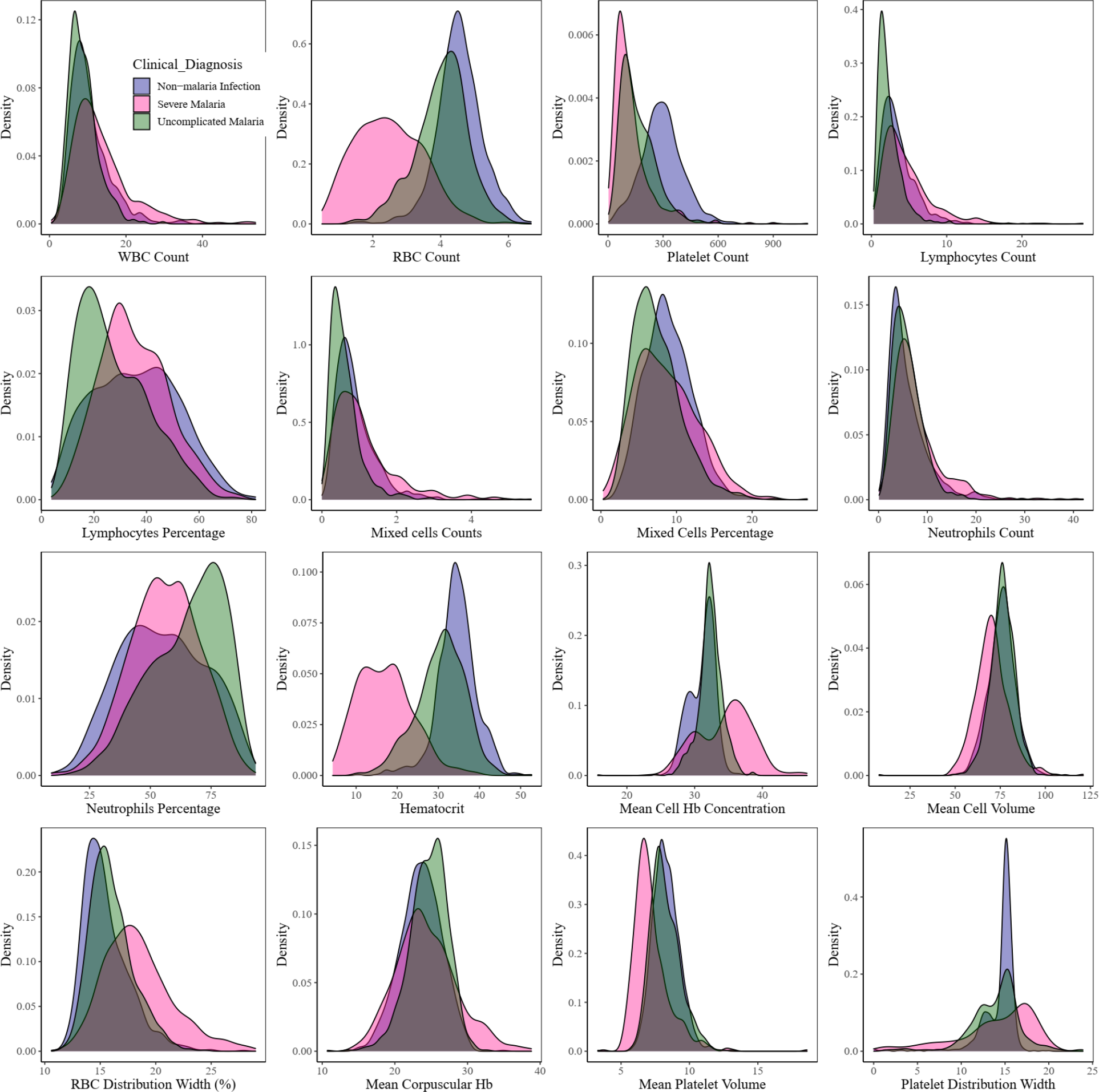
Density estimates of the haematological parameters between nMI, UM and SM cases. A kernel density-plots indicates the distribution of haematological data across each parameter. The plot uses kernel density estimate to plot allowing for smoother distributions by smoothing out the noise. The peaks of each density plot are displaying the help display where values are concentrated over the interval. Each plot is labelled under it the parameter it’s estimating.

To further confirm this hypothesis, the median was used to split the variables into categorical variables (low and high levels). The relationship or predictive value of the categorical parameters to accurately classify the clinical diagnosis was determined using contingency tables (Additional File 1, sheet 2). The percentage number of individuals who had low levels of each parameter and were classified with nMI ranged from 29-70 % (UM group), 7-82 % (SM-group) (Figure 4A). Comparatively, the percentage of individuals who had low levels of each parameter and were classified with UM ranged between 30 - 71 %, while the percentage of individuals who were classified with SM ranged between 17 – 91 % (Figure 4B). There were similar trends for percentage number of individuals who had high levels of each parameter and were classified with either nMI, UM, and SM (Figure 4C –D).

**Figure 4.**
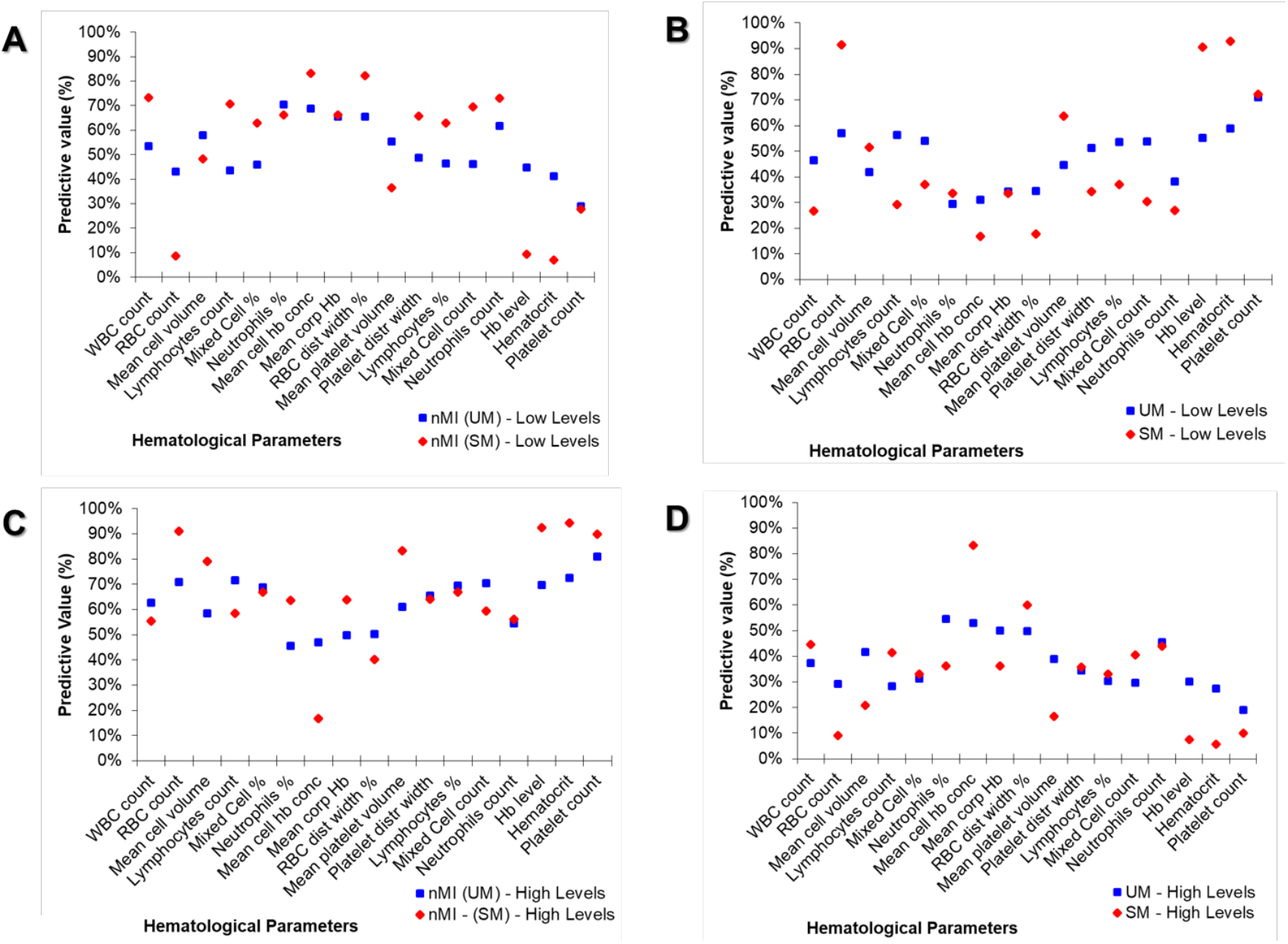
Non-symmetrical predictive values of clinical diagnosis using median cut-off (high vs Low) of the haematological parameters. A “median split” was used to divide each quantitative parameter into categorical variables by the median value (calculated as a mean of nMI and UM or SM median value shown in Table 2). The predictive values are calculated from contingency tables (Supplementary Table 2). (A) Shows the percentages of predicting nMI from low levels, (B) percentages of predicting SM or UM using the low levels, (C) percentages of predicting nMI using high levels and (D) predictive values of UM or SM using high levels.

Additionally, we determined whether the levels could predict whether an individual has UM or SM. First, we predicted UM, and majority of the parameters were associated with clinical diagnosis of SM and nMI (*P*<.001), except mean cell volume, mixed cells counts and neutrophil counts. The parameters that were not associated for nMI-UM category were WBC and RBC counts, Mixed cells percentage, neutrophil percentages, mean cell Hb concentration, mean platelet volume and neutrophil counts (Supplementary Table 2). Furthermore, some of the haematological parameters had a 95 % confidence interval that included the null value (1) when evaluating the odds ratios, which signifies that they are not significantly associated with clinical diagnosis (Supplementary Table 2).

### Machine learning attained over 77.7 % accuracy in classifying clinical malaria from nMI

Since there is no clear distinction between the distributions, and the inability of the median based categories to clearly classify the participant’s clinical diagnosis, we sought to evaluate six ML approaches to classify clinical malaria from nMI. The UM vs nMI model was trained on 942 samples, validated on 403 samples, and tested on 336 samples for each ML approach. The SM vs nMI model was trained on 843 samples, validated on 361 samples, and tested on 300 samples for each ML approach (Figure 1). Amongst the six ML approaches, the training accuracies ranged between 0.794 – 0.857 to classify UM while the training accuracies ranged between 0.937-0.985 in classifying SM. The test accuracies ranged from 0.777 to 0.857 for the UM model, and 0.930 to 0.973 for SM model (Supplementary Table 3). The SVM approach and the ANN generated the overall best classification outcome.

Hyperparameter tuning (n=55,290 combinations) showed that the optimal model for multi-classification had 0.831 training accuracy with a model capacity of 3 layers (128, 64 and 16), with dropouts of 0.4 for layer 1, 0.3 for layer 2, and 0.4 for layer 3, and learning rate of 0.001. The optimal model (n=55,290 combinations) for ANN (nMI vs SM) with 0.975 accuracy had a model capacity of 3 layers (16, 128, and 256 RELU units respectively), the dropout rate was 0.2 and 0.4 for the first two layers, the last layer had 0.1, and a learning rate of 0.0001. The optimal model (n=55,290 combinations) for ANN (nMI vs UM) with 0.869 training accuracy had a model capacity of 3 hidden layers of 256, 64, and 16 RELU units respectively, the dropout rate was 0.1 for the first and last layer and 0.3 for the second layer, and a learning rate of 0.0001. Training and validation history plots for the ANN showed good levelling off for accuracy and loss, as well as acceptable divergence between training loss/accuracy and validation loss/accuracy for all the three models (Supplementary figure 3).

Also, the history plots suggest that there was near zero over-fitting or under-fitting of the data as indicated by closeness of the training and validation curves (Supplementary figure 3). The ANN (UM vs. nMI), achieved 0.856 training accuracy and 0.842 validation accuracy, while the testing accuracy of the model was 0.801 (Kappa 0.583) (Table 3). The training and testing accuracies demonstrate the confidence of the networks in classifying UM. The ANN (SM vs. nMI) achieved a higher accuracy (≥ 0.960) for training, validation and testing accuracy (Table 3). Both ANN had an F1 score of above 0.747, which means the model can be used for the classification of clinical malaria (Table 3). Since the binary classifiers had the best performance, we also performed a multi-classification analysis to assess the ability of the ANN to differentiate among UM, SM and nMI. The data available for the multi-classification model was 2,207 samples, which were split to 80 % training (n=1,766) and 20 % testing (n=441). The training data was further split to 70 % (n=1,236) training and 30 % (n=530) cross-validation with accuracies of 0.862 and 0.828 respectively. The test accuracy was 0.853 (kappa = 0.768), the precision, recall and F1-score of the model was 0.747 (Table 3). The accuracy of multi-classification model provides confidence in the binary classifications.

**Table 3.** Performance of classification models for identifying parameters that can be classified with clinical malaria. Training and cross-validation accuracy as well as testing accuracy, area under the ROC curve (AUC), Precision, Recall and F1-score. Multiclass analysis among all three-disease conditions, training accuracy was 0.862 with 0.828 validation accuracy. The model classified the three classes with 0.853 test accuracy. The ANN (UM vs nMI) had accuracy of ≥ 0.801 for training, validation and testing accuracy. The ANN (SM vs nMI) had the highest classification accuracy of ≥ 0.960.

| ANN | UM vs SM vs nMI | UM vs nMI | SM vs nMI |
| --- | --- | --- | --- |
| **Model type** | Multi-classification model | Binary model | Binary model |
| **Data splitting** |  |  |  |
| Total data (100%) | n=2207 | n=1681 | n=1504 |
| Training & validation data (80%) | n=1766 | n=1345 | n=1204 |
| Testing data (20%) | n=441 | n=336 | n=300 |
| **Training performance** |  |  |  |
| Training accuracy | 0.862 | 0.856 | 0.985 |
| Training Loss | 0.396 | 0.425 | 0.062 |
| Validation accuracy | 0.828 | 0.842 | 0.978 |
| Validation loss | 0.432 | 0.434 | 0.102 |
| **Testing performance** |  |  |  |
| Testing accuracy | 0.853 | 0.801 | 0.960 |
| Kappa | 0.768 | 0.583 | 0.913 |
| ROC_AUC | NA<sup>a</sup> | 0.866 | 0.983 |
| Precision | 0.855 | 0.780 | 0.971 |
| Recall | 0.856 | 0.717 | 0.918 |
| F1- Score | 0.856 | 0.747 | 0.944 |

<sup>a</sup> We did not generate ROC-AUC for multiclassification models

### Diagnostic value of the models using ROC curves

Having shown the accuracy of the models, we determined the ROC curves of ANN (UM vs nMI) and ANN (SM vs nMI) to show the diagnostic ability of these binary classifiers. Both classifiers had very good performance with an AUC 0.866 for ANN (UM vs. nMI) and AUC of 0.983 for ANN (SM vs. nMI) (Figure 5 and Table 3). This showed that the models could be used to distinguish individuals with SM or UM from those with nMI. The cut-offs for UM show that there is a trade-off in sensitivity and specificity as the cut-off increases or decreases, which is not the case for SM. These results could frame the clinical utility of the models and provide a benchmark for future studies.

**Figure 5.**
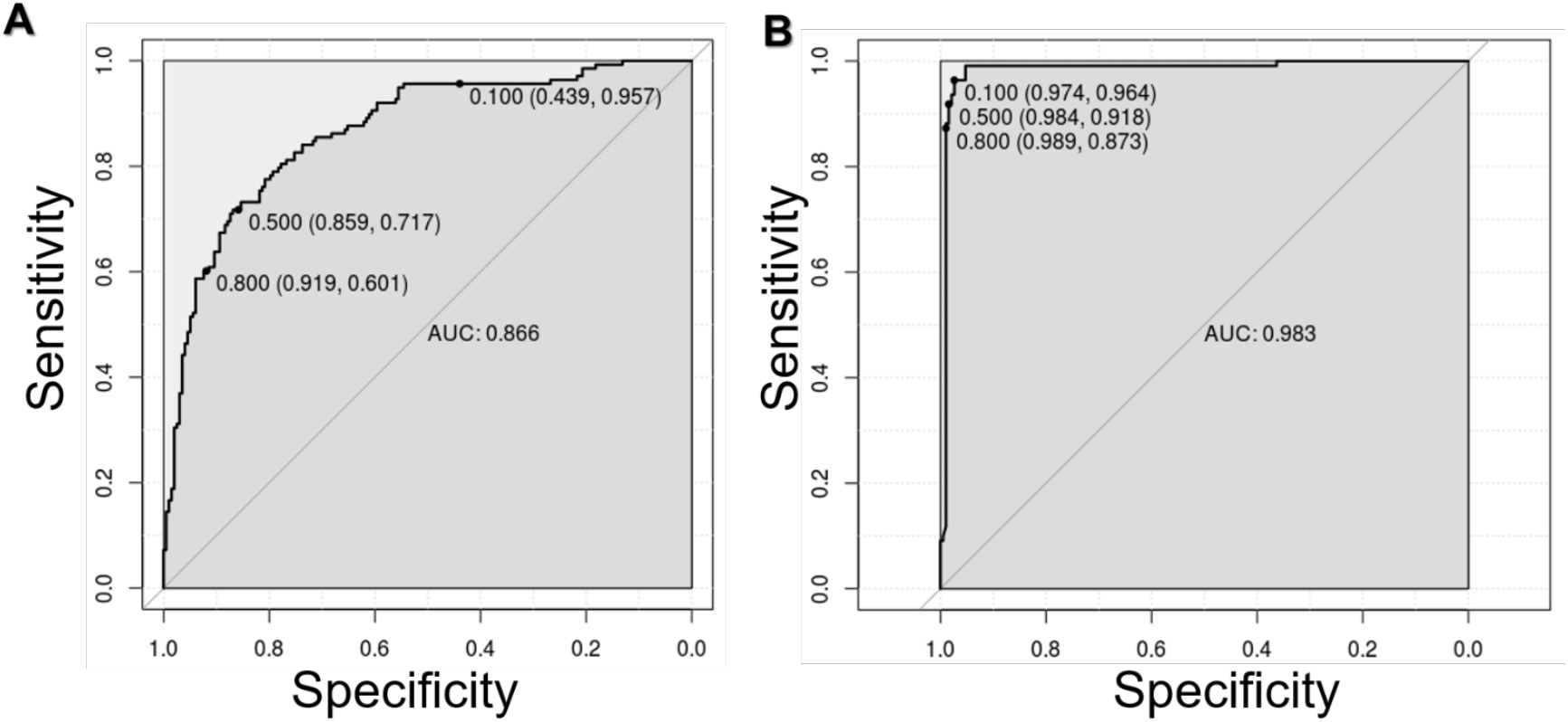
ROC curve for classification of SM was near perfect. The ROC curves plots sensitivity versus specificity for all possible cut offs. Each point on the curve represents a different cut-off value, which are connected to form a curve. The diagonal line is a reference line for the ROC curve. A) ROC for the ANN (UM vs nMI) with an area under the curve (AUC) of 0.866 which is basically an average of true positive rate across all possible false positive rates. B) ROC for the ANN (SM vs nMI) is right angled which means its near perfect with an AUC of 0.983. The levels of AUC indicate a good performance of the models in classifying UM and SM.

### Platelet and RBC counts classify clinical malaria from non-malaria infections

The models were investigated to identify which haematological parameters were classified to be important for either SM, UM or nMI using local interpretable model-agnostic explanations (LIME). Case by case analysis of the individuals showed that some haematological parameters are important classifiers of UM (Supplementary figure 3). Case by case analysis was merged into heatmap to generate a consolidated picture of useful parameters for classification (Figure 6). The top three parameters that had low feature weights for UM are platelet counts, RBC counts and lymphocyte percentages (Figure 6A). Based on the order of importance, the top three parameters that were important for SM classification include RBC counts, platelet counts and mean platelet volume (Figure 6B). This shows that both platelet and RBC counts are important parameters for clinical malaria while the lymphocyte percentages was unique for UM. These parameters might be used to classify clinical malaria cases from nMI, with a very good diagnostic ability as shown by the ROC analysis (Figure 4).

**Figure 6.**
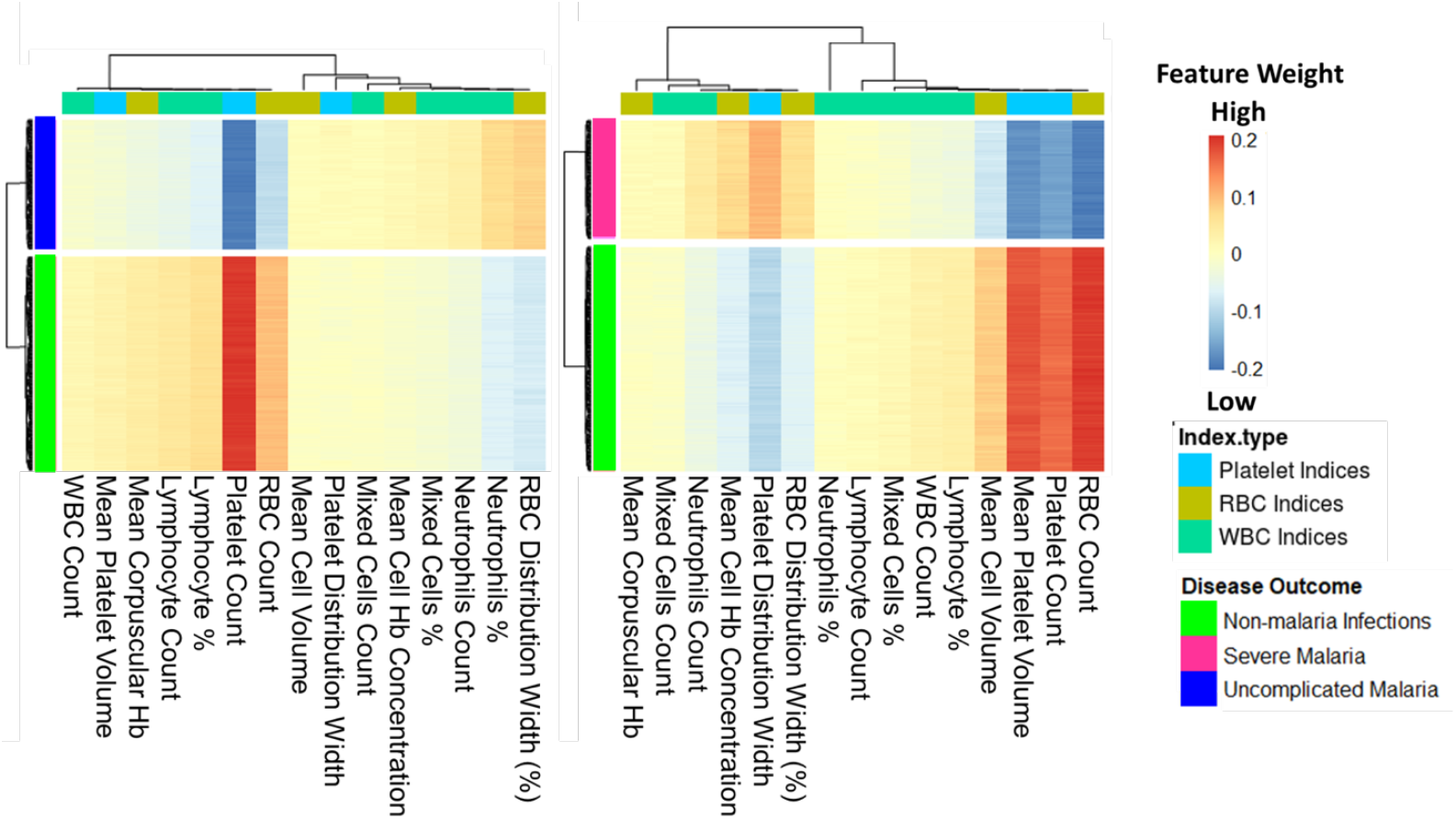
Platelet and RBC counts classified as classifiers of both UM and SM. The Keras model was explained using local interpretable model-agonistic explanations (LIME Package in R-software). The explainer results of the test data, which are represented as feature weights, were extracted from the explainer and used to plot the heatmaps to show a consolidated effect of the parameters. The weights that are < - 0.1 indicate that they are low during UM or SM. **Left** heatmap shows that platelet, RBC, and lymphocytes percentages/counts, can classify UM. **Right** heatmap shows the haematological parameters that can classify SM, and they include RBC counts, mean platelet volume, platelet counts and MCV.

### Patient age and sampling location do not affect the model classifications

We further tested if the models are agnostic to age and location variance. There was a significant difference in patient age between nMI and UM (*P*<.001), but there was no significant difference in samples within Kintampo as well as children under the age of 4 years (Figure 7, supplementary figure 5 & 6). The performance accuracy of the random forest models was 0.806, 0.767, and 0.768 for model 1, 2 and 3 respectively (Figure 7). The most important parameters that featured across the three models were platelet and RBC counts, which are similar to the top two parameters identified by the ANN. Therefore, the data illustrates that age and location do not affect model classifications, and the platelet or RBC counts determined by ANN can be used to reliably classify clinical malaria from nMI in these datasets.

**Figure 7.**
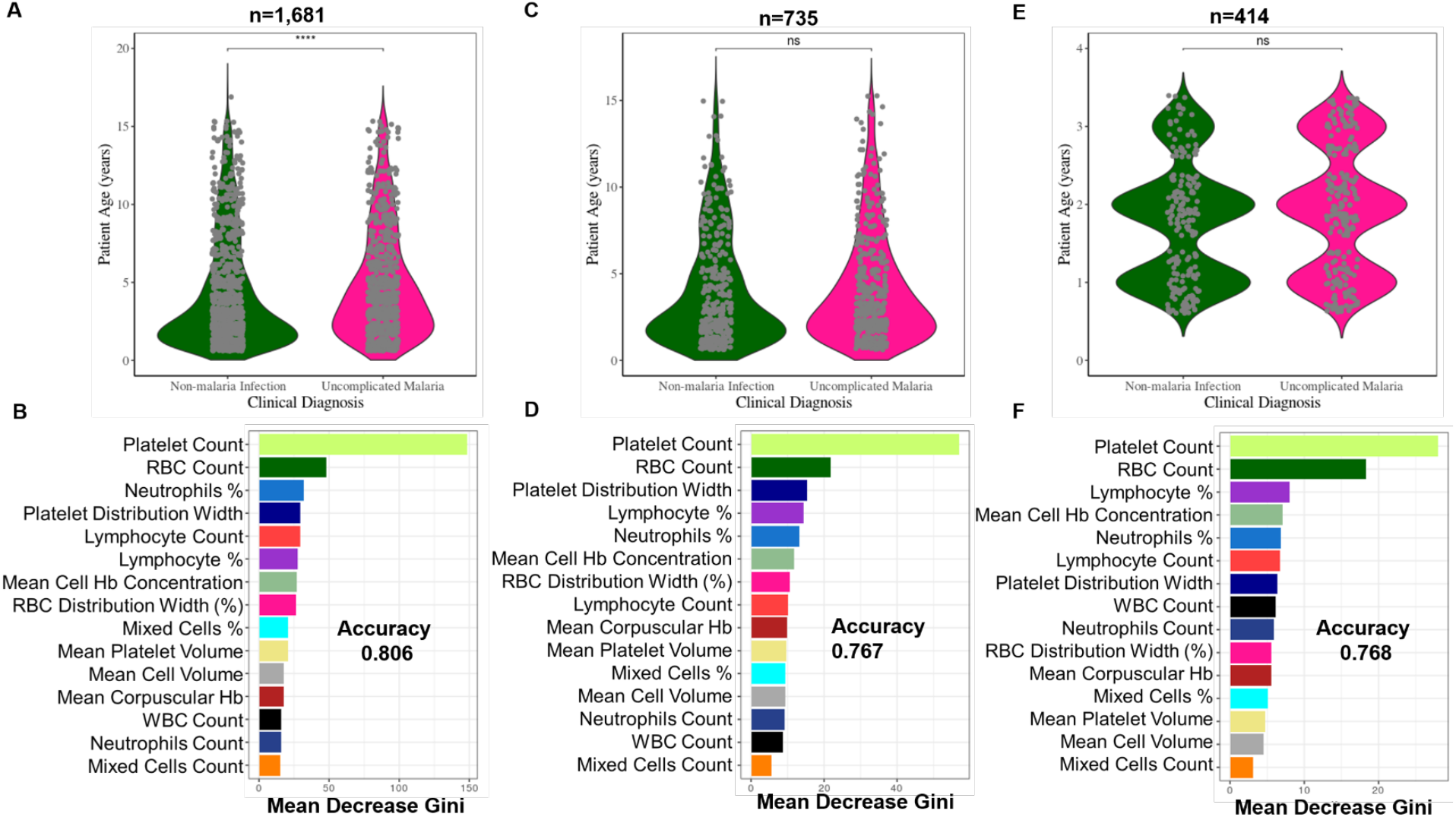
Classification of haematological parameters using random forest shows that patient age and sampling location do not affect the ML models. Three models were generated; (A) a model for all the UM and nMI cases (n=1,681), (B) a model for UM and nMI from Kintampo cases only (n=735), and (C) a model for only Kintampo cases and ages >4 years, whereby there was no significant difference between the nMI and UM (n=414). The samples for each model were split 80% training and 20% for testing. The accuracy of the models was 0.806, 0.767 and 0.768 respectively. The most important feature across the three models was Platelet and RBC counts.

## Discussion

Automated CBC is one of the blood tests routinely performed for children presenting to health facilities with fever. However, CBC analysis generates a significant amount of data on a range of haematological parameters, the data is underutilized with only Hb and Hct levels being routinely used as an indicator of clinical malaria. Thus, an automated algorithm to detect malaria based on the haematological parameters as outlined in this study, could have great value as a complementary malaria diagnostic strategy, particularly at front-line health centers where CBC is routinely done. Such an algorithm also has the added value of enabling the monitoring of treatment outcomes for in-patients.

In malaria endemic settings, malaria rapid diagnostic tests (mRDTs) have revolutionized diagnosis and significantly reduced presumptive treatment, particularly in rural settings where trained microscopists are lacking [3]. However, reports of emerging *Pfhrp2/3* gene deletions threaten the future reliability of the RDTs. False negative RDT results are also known to occur in low density infections [7–9]. Thus, an approach that is automated and agnostic to parasite genetic variation is critical both as a fail-safe and a surveillance strategy for false negative mRDTs (which might occur due to supply chain mismanagement or gene variation) [9]. In very low transmission settings, ML models have the potential to replace the primary use of mRDTs when diagnostic yield of mRDTs becomes very low (i.e. many mRDTs needed to detect a single case of malaria). In non-endemic settings where malaria may occur in immigrants and non-immune travelers, the models may allow another fail-safe mechanism in case the diagnosis of malaria was not suspected by clinicians and malaria RDT or microscopy was not performed. Despite these advantages, there would be a little extra cost associated with incorporating the algorithm and an automated message into haematology analyzer output, a message that can prompt clinicians to consider malaria in the presence of suggestive haematological features.

Previous ML studies have looked into haematological parameters more generally and to classify sickle cell anaemia using deep convolutional networks [43, 44], but did not classify clinical diagnosis. For the first time, ML approaches that can classify infections in children based on haematological parameters have been generated. Six different ML methods were evaluated and they were all shown to classify clinical malaria from nMI with high accuracy especially the SVM and the ANN. We used the ANN to deconvolute the results: it identified platelet and RBC counts as the top features in classifying both UM or SM from nMI. Low RBC counts can be attributed to extensively parasitized RBCs, which are sequestered during SM [45]. This highlights the significance of RBC counts during *Plasmodium falciparum* malaria infections. In most occasions except cerebral malaria, SM is associated with anemia due to RBC lysis during parasite invasion as well as many other RBC abnormalities [46]. This makes the diagnosis of SM much easier than UM, whereby one parameter, such as Hb level of <5g/dL, can diagnose or classify the disease.

Cohen et al, analyzed data from 680,964 individuals with fever and confirmed that majority of antimalarial drugs are given to malaria negative individuals [47]. Overtreatment indicates that, most nMI can go without being treated, for their true cause, which is also possible for UM and this can lead to drug resistance. Therefore, the difference between febrile outpatient infections is far more challenging, especially between nMI and UM due to similarity in clinical presentations. In large population studies, differences in median can be significant but they do not necessarily distinguish the populations as either nMI or UM as observed in this study. But, using the ML approach shown here, distinguishing the nMI and UM can be improved by combining all haematological parameters and learning the data-patterns before making classifications. The predictions made by ML are more accurate and reliable, and can be improved by analyzing more datasets. Lymphocytes counts/percentage were identified to be affected during UM, and can be used to distinguish UM from nMI, mainly because individuals with malaria generally have a distinct immune response compared to nMI individuals [29, 48, 49].

Previous work in our laboratory showed differences in haematological presentation among areas of varying transmission intensity in Ghana [50]. To show that differences in age and transmission zones (sampling location) are not driving our diagnostic classifications, we down-sampled the data and used random forest to perform the classifications. The results showed that, platelet and RBC counts were the key features in classifying UM and nMI regardless of age and sampling location of the participants. There were differences in the top three important features between the random forest and ANN, but this could be due to the differences in approach of each algorithm [25, 51]. This illustrates that the patient age and location do not substantially influence the diagnostic classifications. The ROC curves further showed that the models could be used for diagnosis with very reliable AUC values.

There are four limitations to be considered in the use of this ML approach in routine diagnosis and the generalization of our approach. First, the models can distinguish between nMI and clinical malaria, but whether they can be used to distinguish the clinical disease state will depend on the pre-test probability or prevalence of malaria in different settings. Second, all study subjects being Ghanaian children may limit the generalizability of the models; this is also the case for the limited range of SM manifestations in our dataset, and the spectrum of laboratory confirmed nMI. Lastly, the study did not have adults >15 years to comparatively understand the role of age in differentiating clinical malaria based on haematological parameters. Therefore, we recommend that more studies are needed to inform the broader utility of this work. Despite that only 4.6 % (75/1,645) of the cases were discordant between microscopy and RDT, probably due to *hrp2/hrp3* deletions, there is an insignificant chance that misclassification of malaria could have had an impact on our study. These limitations will be taken into account for further studies to inform the broader clinical utility of this work.

## Conclusions

Fever is the most common symptom reported in sSA, and correct diagnosis of the implicated pathogen is of high importance for precision medicine. Personalized treatment reduces overtreatment, decreases malaria mortality and antimalarial resistance. This report demonstrates proof-of-principle that ML can be used to distinguish clinical malaria from nMI using routine haematological data. Case by case analysis showed that, the models can make classifications based on combination of three parameters: platelet and RBC counts, lymphocytes counts/percentage and mean platelet volume. These could be used for precision diagnosis of an individual’s risk of having malaria, to inform the need for confirmatory diagnosis by microscopy or to prompt investigation for other diagnoses when malaria is unlikely. Further work is to calibrate and improve the classification capability of the model using more data from other geographical and transmission settings, demographic groups, co-infections, and different disease severities. Our findings hold promise for the design of clinical software to support diagnosis of malaria in the WHO African region, and might also prove useful for diagnosis of malaria in returning travelers from non-endemic countries.

## Data Availability

All data will be made available on acceptance of paper.

## Acknowledgments

We acknowledge the study participants as well as the staff of Kintampo Municipal, who provided support for this study. We are grateful to Emmanual Allotey, Prince Nyarko, Henrietta Mensa-Brown, Felix Ansah, Jersley Chirawurah, Jonas Kengne, and Nancy Nyakoe for their contributions on the data collection and constructive criticism of the work. We also express our sincere gratitude to Dorothy Annan, Deborah Mettle, Bright Yemi, Rachel Abban, Samirah Saiid, Joyceline Kwarko, and Israel Osei for assisting in cross-referencing the data. We acknowledge University of Ghana for providing the high performance computing resources (the ZUPUTO) used for this work.

## Author’s Contributions

CM, GA, LA, TO, SB, DA, AC, and VA contributed to design and conceptualization of the work, as well as editing and critique of the manuscript drafts. LA, JA, DA, AO, and NA contributed to data collection. CM performed data analysis, including building the models, model interpretation and drafting the manuscript. All authors read and approved the final manuscript.

## Funding

The study was supported by a DELTAS Africa grant (DEL-15-007: Awandare). The DELTAS Africa Initiative is an independent funding scheme of the African Academy of Sciences (AAS)’s Alliance for Accelerating Excellence in Science in Africa (AESA) and supported by the New Partnership for Africa’s Development Planning and Coordinating Agency (NEPAD Agency) with funding from the Wellcome Trust (107755/Z/15/Z: Awandare) and the UK government. The funder had no role in study design and where to publish. The views expressed in this publication are those of the author(s) and not necessarily those of AAS, NEPAD Agency, Wellcome Trust or the UK government.

## Availability of the data

The dataset supporting the conclusions of this article is included within the article and its additional files.

## Ethics approval and consent to participate

Ethical clearance was obtained from Ghana Health Service Ethics Review Committee and Noguchi Memorial Institute of Medical Research Institutional Review Board. All participants were provided with written informed consent prior to inclusion in the study.

## Consent for Publication

All authors gave consent for publication

## Competing Interests

The authors declare that they have no competing interests.

## Supplementary Tables

**Supplementary Table 1.** The list of haematological parameters adopted from laboratory procedure manual by the CDC [52]. The table outlines the description of the parameters, abbreviation, mode of measurement, pulse size, and the units of reporting.

| Cell | Parameter | Measured | Pulse Size | Reported Units |
| --- | --- | --- | --- | --- |
| WBC | **White Blood Cell Count**<br>This is the number of leukocytes measured directly, multiplied by the calibration constant, and expressed as $n \times 10^3$ cells/μL | WBC bath | $\geq 35$ fL | $n \times 10^3$ cells/μL |
| RBC | **Red Blood Cell Count**<br>This is the number of erythrocytes measured directly, multiplied by the calibration constant, and expressed as $n \times 10^6$ cells/μL | RBC bath | 36 to 360 fL | $n \times 10^6$ cells/μL |
| Hb | **Hemoglobin Concentration**<br>Weight (mass) of hemoglobin determined from the degree of absorbance found through photocurrent transmittance is: $Hb \text{ (g/dL)} = \text{Constant} \times \log_{10}(\text{Reference \% T} / \text{Sample \% T})$ | WBC bath | 525 nm | g/dL |
| Hct | **Hematocrit**<br>This is the relative volume of packed erythrocytes to whole blood, computed as: $Hct (\%) = RBC \times MCV/10$ | Computed | $RBC \times MCV/10$ | % Percent |
| MCV | **Mean Cell Volume**<br>This is the average volume of individual erythrocytes derived from the RBC histogram. | Derived from RBC histogram | # x size of RBC/ Total RBC | fL |
| MCH | **Mean Cell Hemoglobin**<br>This is the weight of hemoglobin in the average erythrocyte count, computed as: $Hb / RBC \times 10$ | Computed | $Hb/RBC \times 10$ | pg |
| MCHC | **Mean Cell Hemoglobin Concentration**<br>This is the average weight of hemoglobin in a measured dilution, computed as: $Hb / Hct \times 100$ | Computed | $Hb/Hct \times 100$ | g/dL |
| RDW | **Red Cell Distribution Width**<br>RDW represents the size distribution spread of the erythrocyte population derived from the RBC histogram. It is the coefficient of variation (CV), expressed in percent, of the RBC size distribution. | Derived from RBC histogram | CV expressed in % of the RBC size distribution | % Percent |
| Plt | **Platelet Count**<br>This is the number of thrombocytes derived from the Plt histogram and multiplied by a calibration constant. This number is expressed as: $n \times 10^3$ cells/μL | RBC bath | 2 to 20 fL | $n \times 10^3$ cells/μL |
| MPV | **Mean Platelet Volume**<br>MPV is the average volume of individual platelets derived from the Plt histogram. It represents the mean volume of the Plt population under the fitted | Derived from Plt histogram | Mean volume of Plt population under the fitted curve x constant | fL |
|  | Plt curve multiplied by a calibration constant, and expressed in femtoliters. |  |  |  |
| NE% | **Neutrophil Percent**<br>The percentages of leukocytes from each category are derived from the scatterplot. | Derived from scatterplot | # cells inside NE area/# cells inside total cell area x 100 | % Percent |
| NE # | **Neutrophil Number**<br>The absolute numbers of leukocytes in each category are computed from the WBC count and the differential percentage parameters. | Absolute number | NE%/100 x WBC Count | 10<sup>3</sup> cells/μL |
| LY% | **Lymphocyte Percent**<br>The percentages of leukocytes from each category are derived from the scatterplot. | Derived from scatterplot | # cells inside LY area/# cells inside total cell area x 100 | % Percent |
| LY# | **Lymphocyte Number**<br>The absolute numbers of leukocytes in each category are computed from the WBC count and the differential percentage parameters. | Absolute number | Ly%/100 x WBC Count | 10<sup>3</sup> cells/μL |
| MO% | **Monocyte Percent**<br>The percentages of leukocytes from each category are derived from the scatterplot. | Derived from scatterplot | # cells inside MO area/# cells inside total cell area x 100 | % Percent |
| MO# | **Monocyte Number**<br>The absolute numbers of leukocytes in each category are computed from the WBC count and the differential percentage parameters. | Absolute number | MO%/100 x WBC Count | 10<sup>3</sup> cells/μL |

\*PDW - Platelet Distribution Width and Pct - Plateletcrit are NOT for diagnostic use and do not print. Coulter uses the value for
PDW is an internal check on the reported platelet parameters

**Supplementary Table 2.**
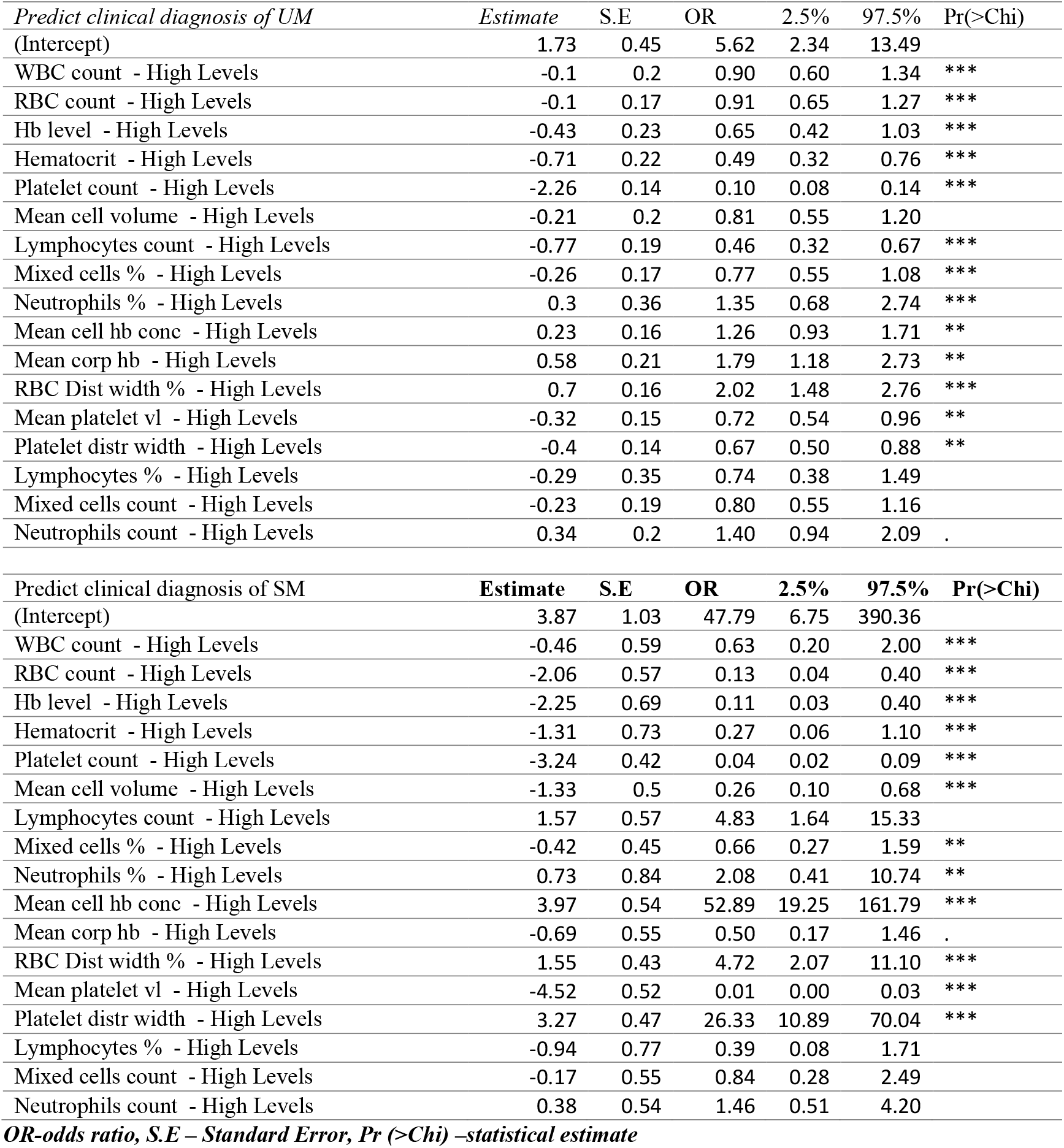
The odds ratio of median categories providing the odd of being diagnosed with either nMI, UM, and SM. The median categories were; low and high levels.

**Supplementary table 3.**
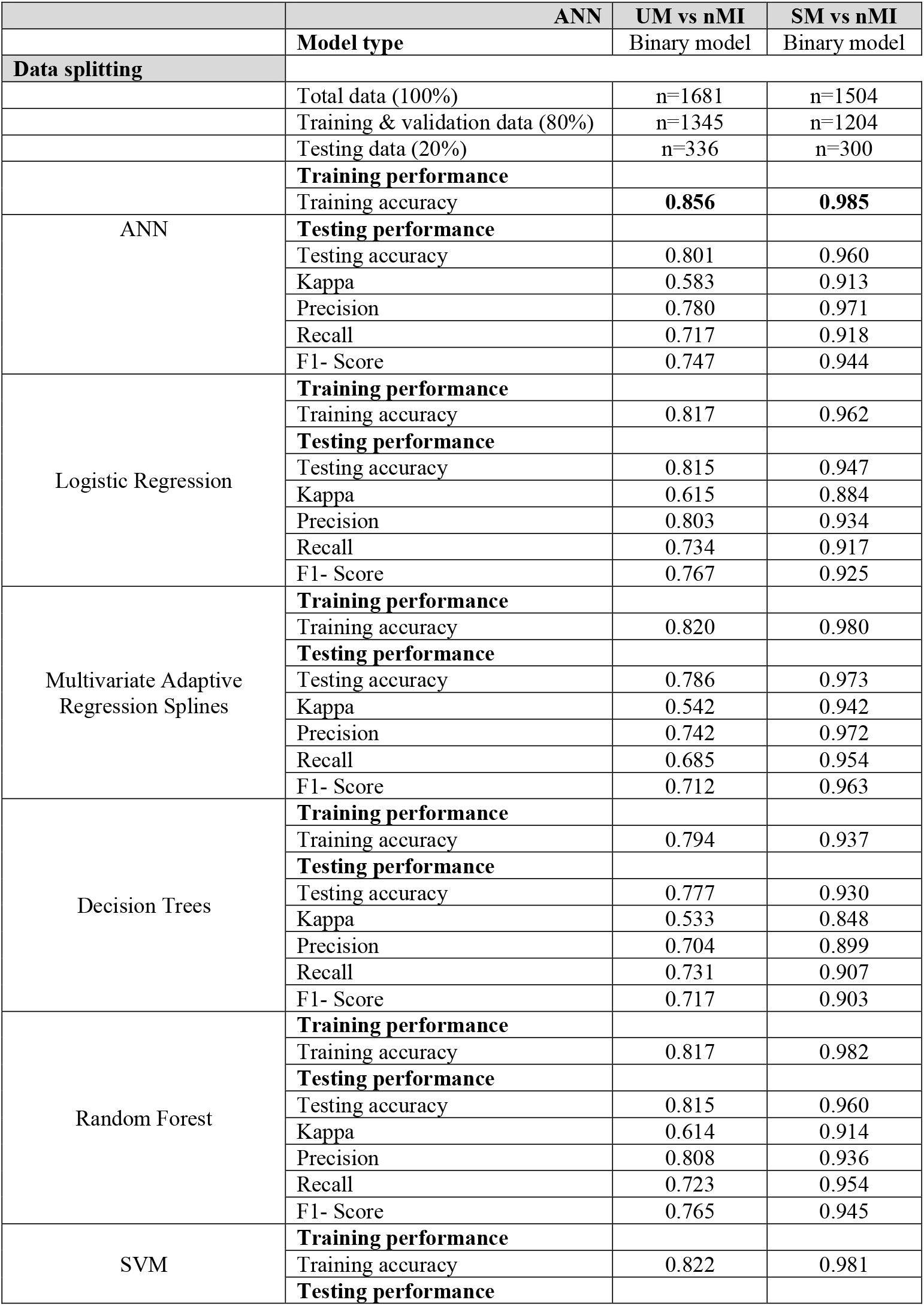
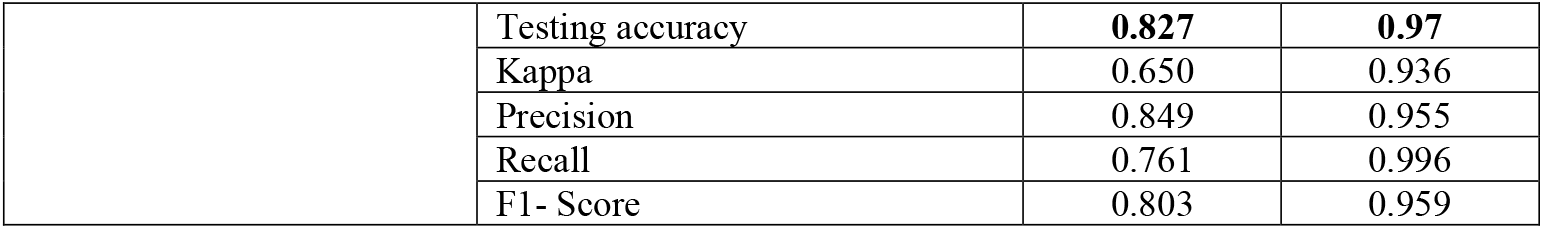
Performance evaluation of six machine learning models to classify clinical malaria outcomes.

**Supplementary figure 1.**
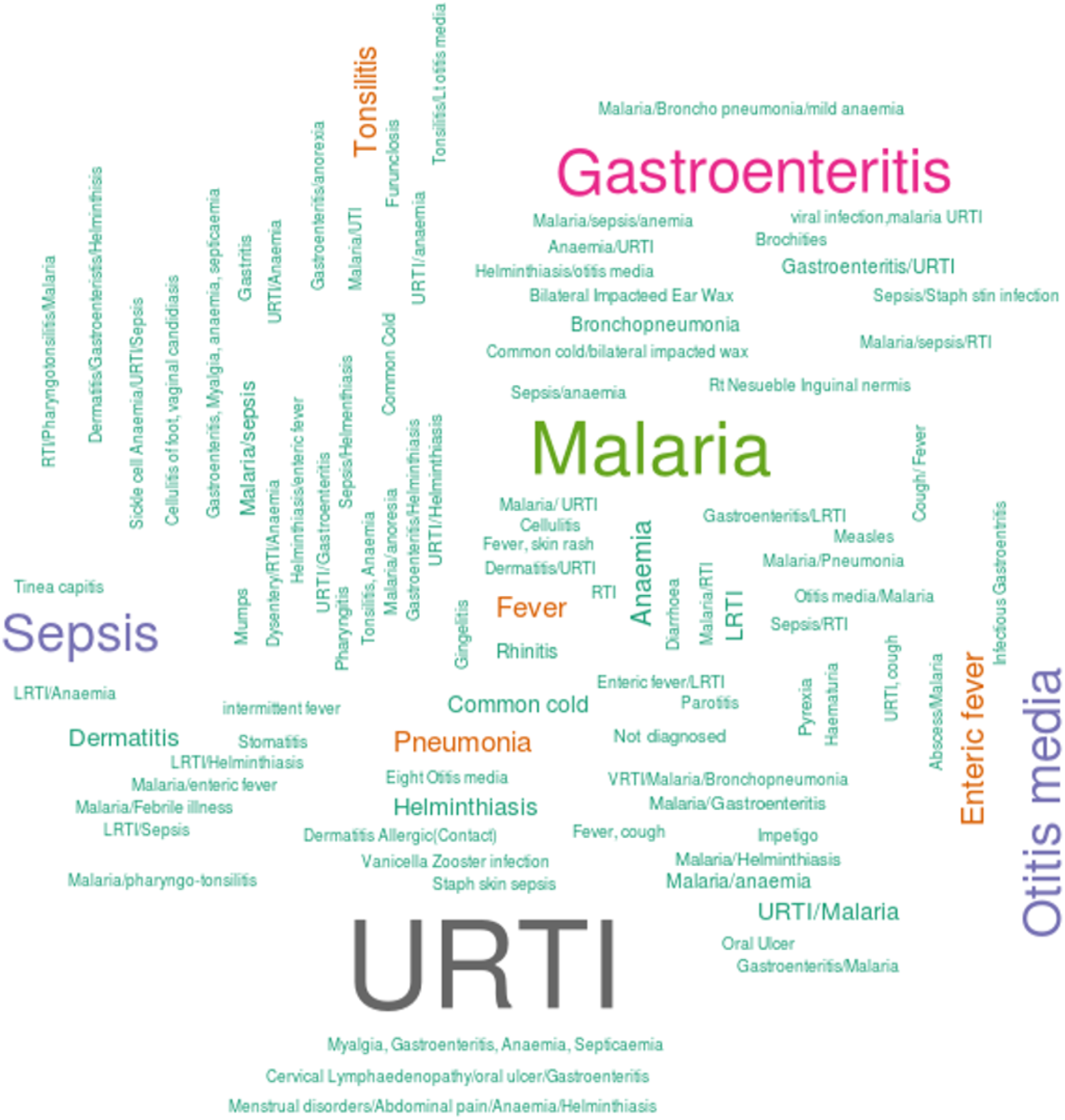
Word cloud of clinical manifestations using clinicians/doctors notes or suspected infections. Top 150 infections that were reported; with majority of the people having upper respiratory tract infections (URTI), followed by Malaria, Gastroenteritis, Sepsis, Otitis media, and Fever. The rest of the diagnosis had a frequency less than 2.

**Supplementary figure 2.**
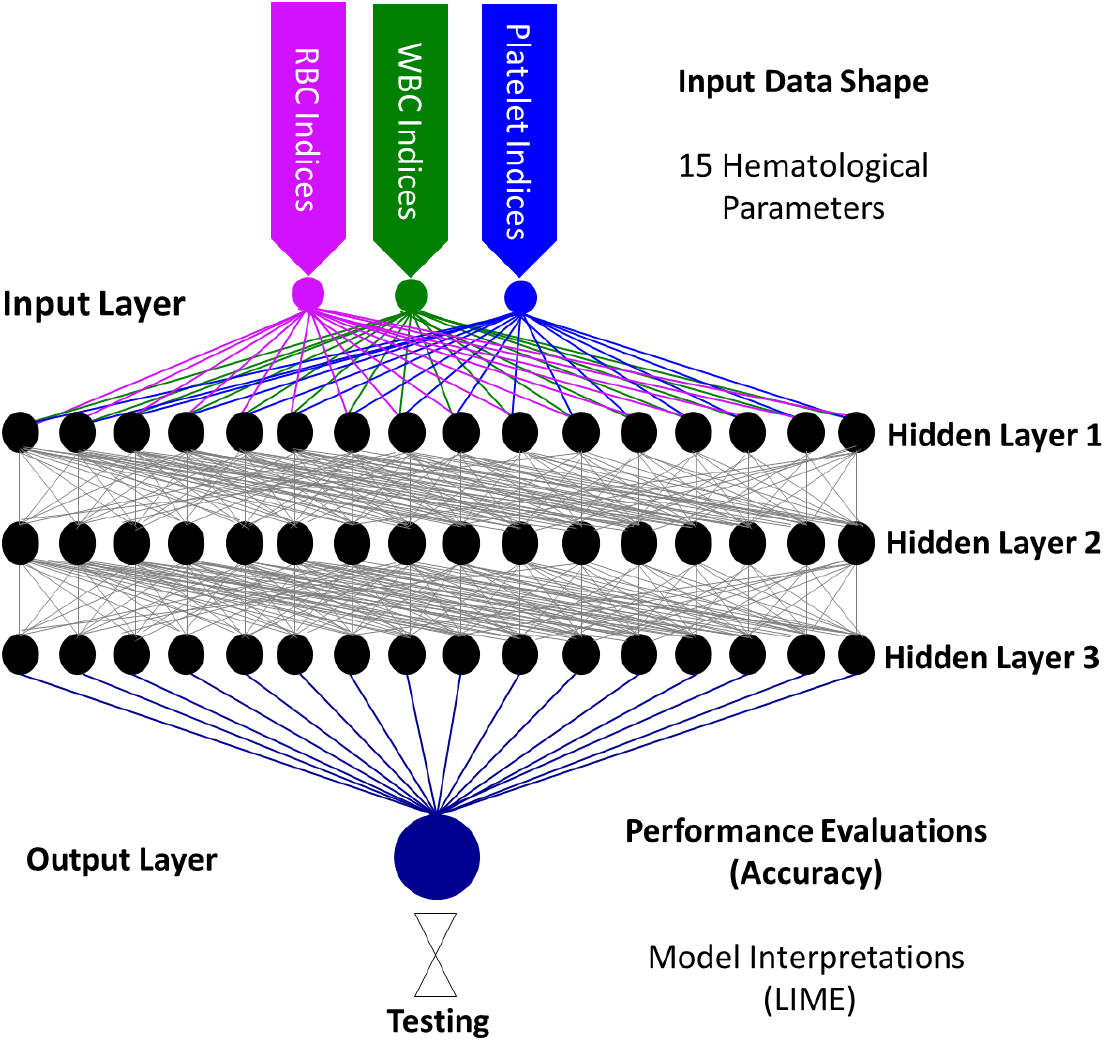
Artificial Neural Network Schematic. Keras Model that is composed of a linear stack of input layers, three hidden layers and one output layer. The input layer composed of an input shape of 15 haematological parameters (RBC parameters, WBC parameters, and Platelet indices). The hidden layers are each composed of a represetative16-stacked unit with ReLU as the activation function. The output layer had sigmoid function as the activation function and uniform initialization. Interpretations of the model classifications were made using local interpretable model-agonistic explanations (LIME Package in R) [41].

**Supplementary figure 3.**
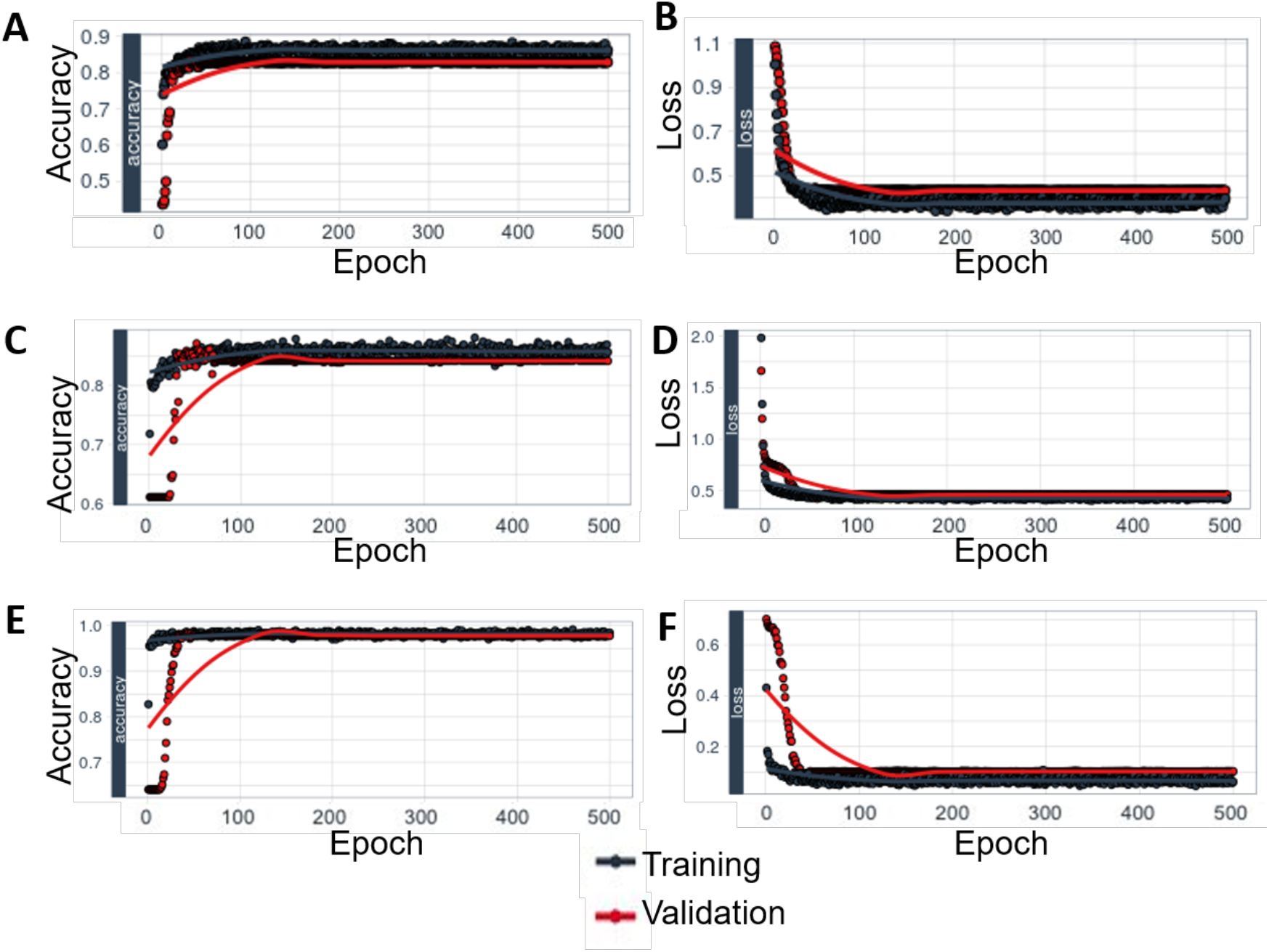
Plot for the training and validation history of the ANN. The figure indicates training and validation history of the model, which shows how accuracy and loss are leveling off, as well as the divergence between training and validation accuracy and training and validation loss. (A and B) Multi-classification of (SM vs. UM vs. nMI) accuracy and loss respectively. (C and D) Accuracy and loss respectively for ANN (UM vs. nMI) binary classifier. (E and F) Accuracy and loss respectively for the ANN (SM vs. nMI) binary classifier. The plots show a minimal model gap between training and validation.

**Supplementary figure 4.**
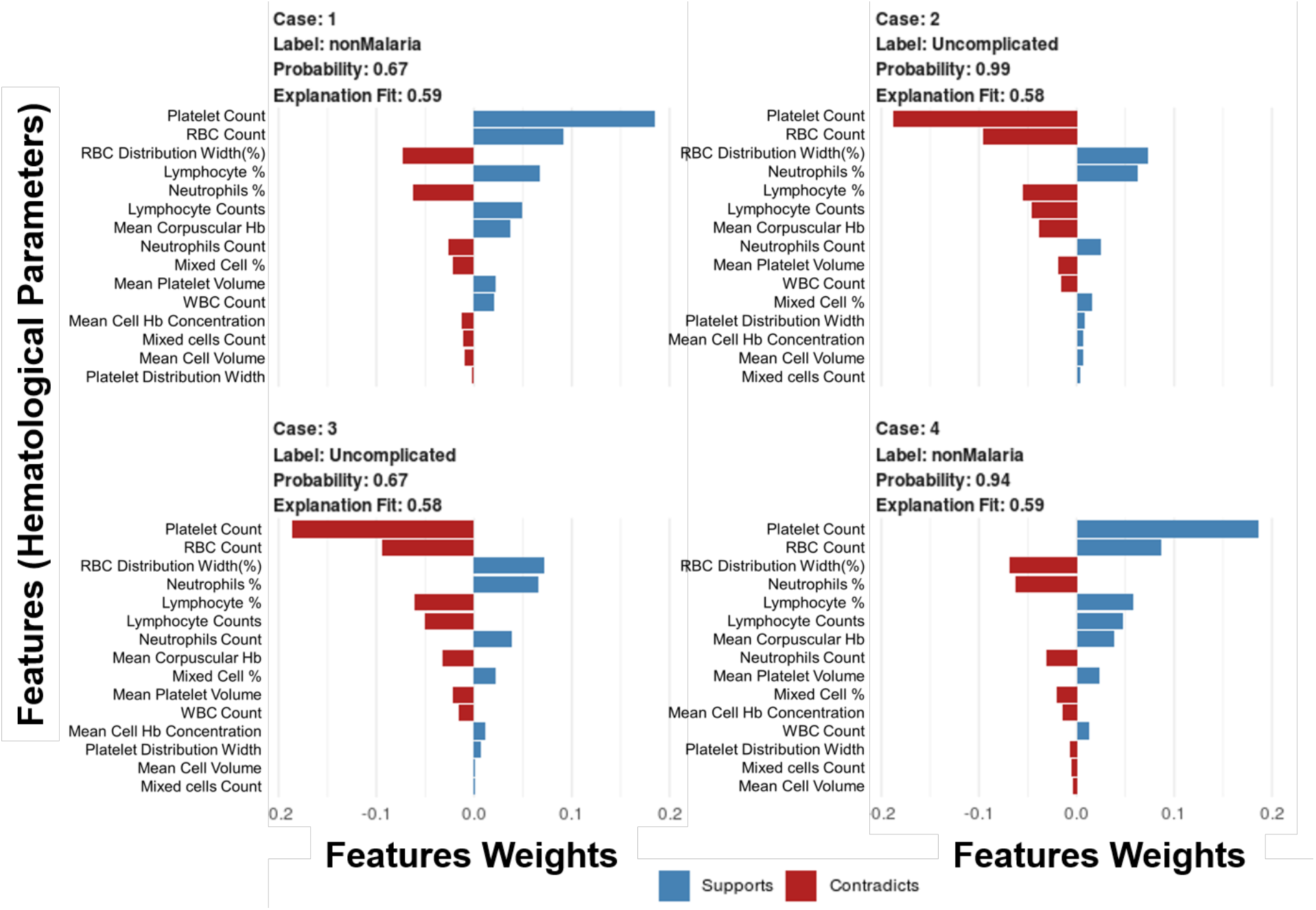
Case by case analysis of the classification capability of the ML models. Samples of four cases in the test dataset were selected to indicate the predictions for each case. Case 1 and 4 are nMI patients, while case 2 and 3 are UM patients. The bars indicate the feature weights for each haematological parameter and whether it is predictive of malaria (supports) or not (contradicts). This figure highlights how the parameters can be used for precision medicine.

**Supplementary figure 5.**
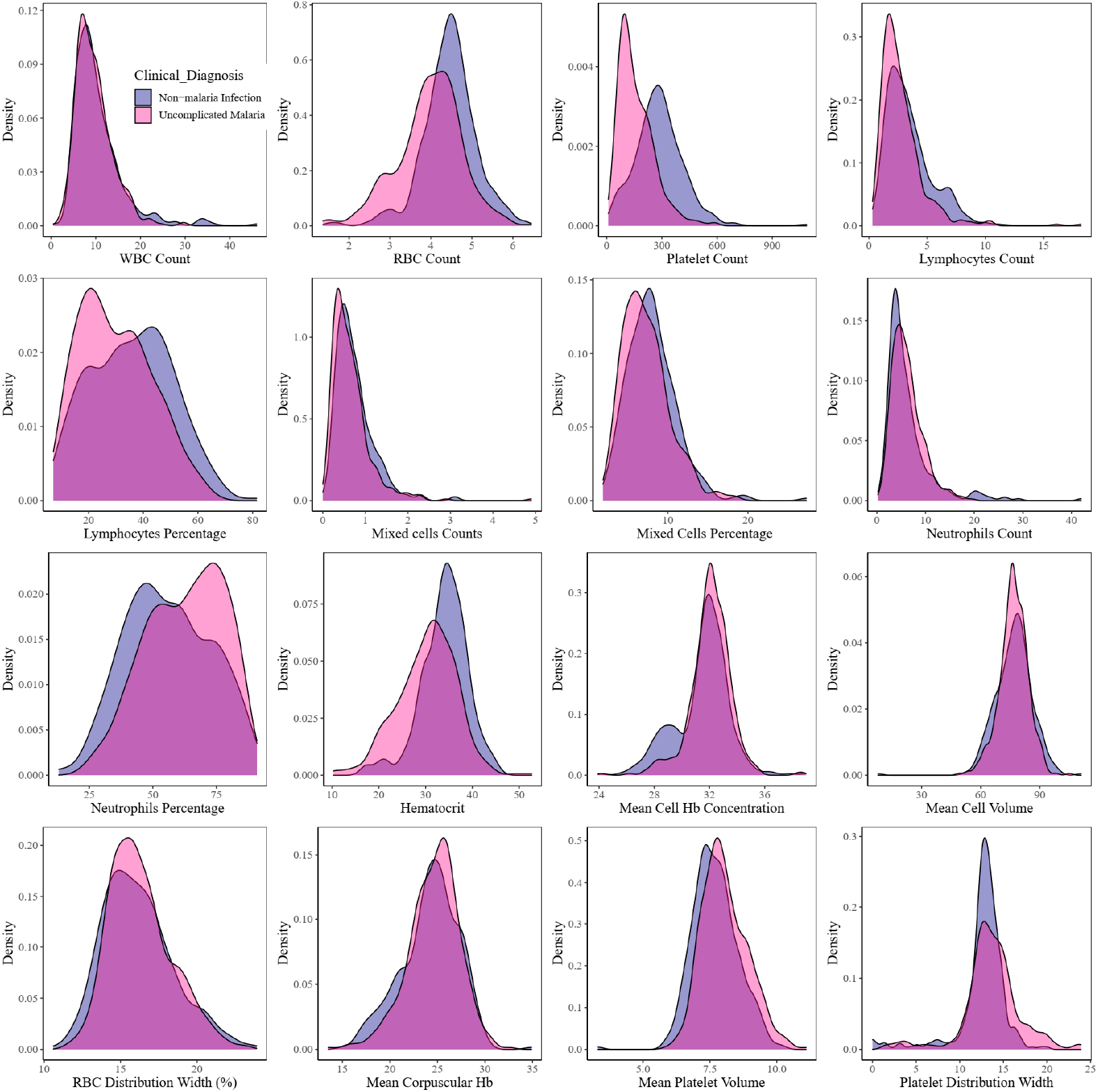
Density estimates of the haematological parameters between nMI and UM cases for sub-sampled data from Kintampo only.

**Supplementary figure 6.**
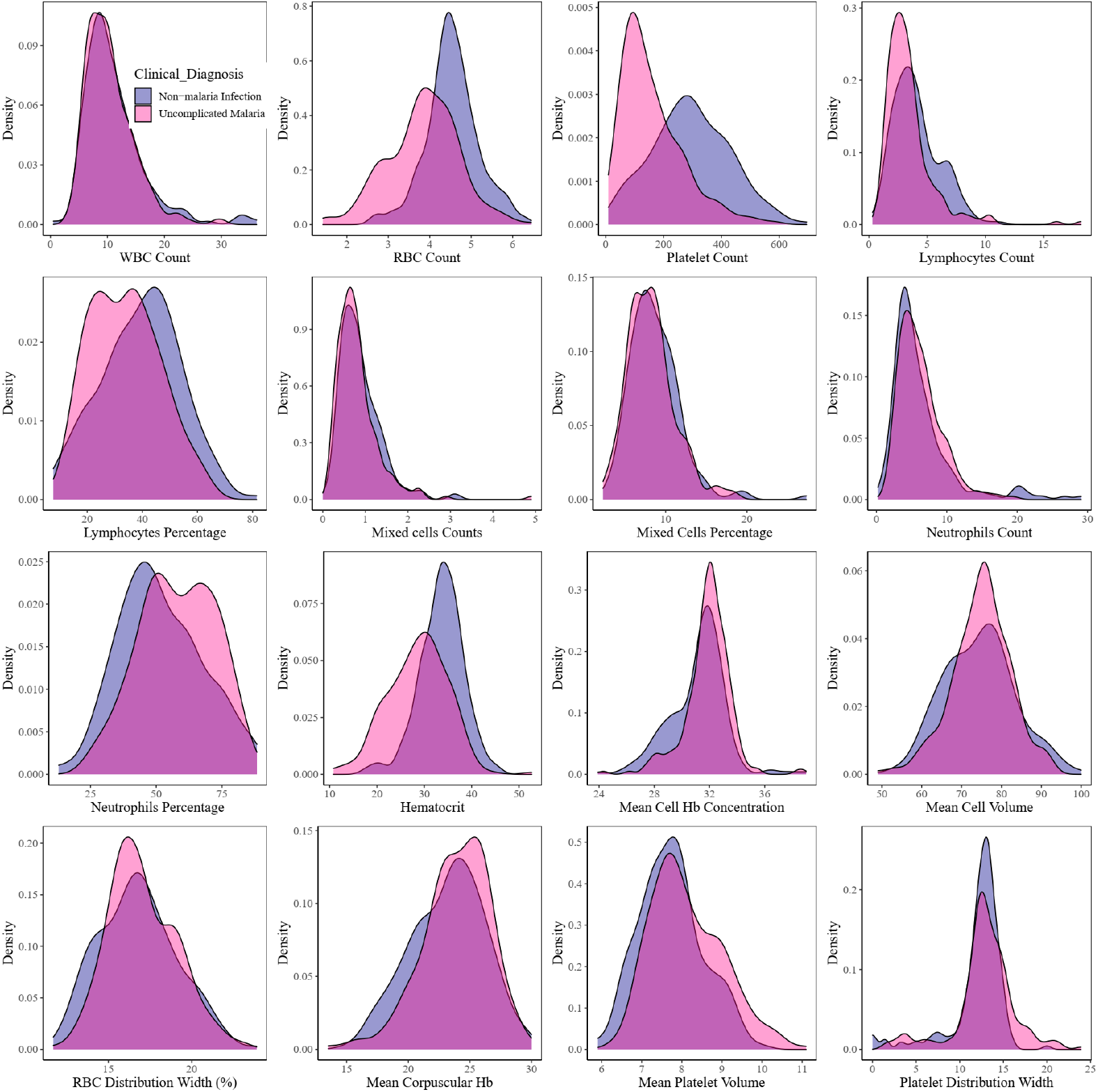
Density estimates of the haematological parameters between nMI, and UM cases for sub-sampled data from Kintampo only, as well limit of children under 4 years of age.

## Notes

### Competing Interest Statement

The authors have declared no competing interest.

